# Infectious SARS-CoV-2 in Exhaled Aerosols and Efficacy of Masks During Early Mild Infection

**DOI:** 10.1101/2021.08.13.21261989

**Authors:** Oluwasanmi O. Adenaiye, Jianyu Lai, P. Jacob Bueno de Mesquita, Filbert Hong, Somayeh Youssefi, Jennifer German, S.-H. Sheldon Tai, Barbara Albert, Maria Schanz, Stuart Weston, Jun Hang, Christian Fung, Hye Kyung Chung, Kristen K. Coleman, Nicolae Sapoval, Todd Treangen, Irina Maljkovic Berry, Kristin Mullins, Matthew Frieman, Tianzhou Ma, Donald K. Milton, for the University of Maryland StopCOVID Research Group

**Author notes:** Dr. Donald K. Milton, University of Maryland School of Public Health, 4200 Valley Drive, College Park, MD 20742; (Alternate: Dr. Filbert Hong, at the same address).

## Abstract

**Background:** SARS-CoV-2 epidemiology implicates airborne transmission; aerosol infectiousness and impacts of masks and variants on aerosol shedding are not well understood.

**Methods:** We recruited COVID-19 cases to give blood, saliva, mid-turbinate and fomite (phone) swabs, and 30-minute breath samples while vocalizing into a Gesundheit-II, with and without masks at up to two visits two days apart. We quantified and sequenced viral RNA, cultured virus, and assayed sera for anti-spike and anti-receptor binding domain antibodies.

**Results:** We enrolled 49 seronegative cases (mean days post onset 3.8 ±2.1), May 2020 through April 2021. We detected SARS-CoV-2 RNA in 45% of fine (≤5 µm), 31% of coarse (>5 µm) aerosols, and 65% of fomite samples overall and in all samples from four alpha-variant cases. Masks reduced viral RNA by 48% (95% confidence interval [CI], 3 to 72%) in fine and by 77% (95% CI, 51 to 89%) in coarse aerosols; cloth and surgical masks were not significantly different. The alpha variant was associated with a 43-fold (95% CI, 6.6 to 280-fold) increase in fine aerosol viral RNA, compared with earlier viruses, that remained a significant 18-fold (95% CI, 3.4 to 92-fold) increase adjusting for viral RNA in saliva, swabs, and other potential confounders. Two fine aerosol samples, collected while participants wore masks, were culture-positive.

**Conclusion:** SARS-CoV-2 is evolving toward more efficient aerosol generation and loose-fitting masks provide significant but only modest source control. Therefore, until vaccination rates are very high, continued layered controls and tight-fitting masks and respirators will be necessary.

**Key Points:**

- Cases exhale infectious viral aerosols
- SARS-CoV-2 evolution favors more efficient aerosol generation.
- Loose-fitting masks moderately reduce viral RNA aerosol.
- Ventilation, filtration, UV air sanitation, and tight-fitting masks are needed to protect vulnerable people in public-facing jobs and indoor spaces.

## Introduction

The World Health Organization[1] and U.S. Centers for Disease Control and Prevention[2] recently acknowledged the growing scientific consensus that inhalation exposure is an important route of SARS-CoV-2 transmission[3,4]. The totality of evidence from epidemiologic and outbreak investigations, combined with data on the size distribution of exhaled aerosols and corresponding quantitative models, is compelling[4]. However, culture of the virus from exhaled aerosols, and direct measures of the efficacy of face masks as source control when worn by actual patients have been lacking. Previous reports of infectious virus[5,6] and viral RNA concentrations in room air[7,8] do not provide a clear picture of how much virus infected persons shed into the air. These gaps lead to uncertainty in estimates of exposure, derived from retrospective analysis of outbreaks[9]. New variants also appear more transmissible, but more quantitative data is needed to discern what that means for implementing effective non-pharmaceutical interventions – still a mainstay of infection protection.

We collected exhaled breath aerosol samples from polymerase chain reaction (PCR)-confirmed COVID-19 cases infected with SARS-CoV-2, including alpha and earlier variants, circulating in a university campus community using a Gesundheit-II (G-II) exhaled breath sampler[10,11]. We measured concentrations of SARS-CoV-2 RNA and recovered infectious virus from respiratory swabs, saliva, and aerosols, analyzed the effectiveness of face masks as source control, and examined the impact of the alpha variant on aerosol shedding.

## Materials and Methods

This study was approved by the University of Maryland Institutional Review Board and the Human Research Protection Office of the Department of the Navy. Electronically signed informed consent was obtained and questionnaire data were collected and stored with REDCap[12].

We recruited participants with active infection (defined as positive qRT-PCR for SARS-CoV-2 in respiratory swab or saliva samples) May 2020 through April 2021 from the University of Maryland College Park campus and community though a) daily symptom reporting and weekly pooled saliva testing cohort of 238 volunteers, b) direct recruiting of recently diagnosed cases from local clinics and campus, and c) frequent testing of close contacts of cases for two weeks following last contact. Cases and contacts completed online consents and a questionnaire (see Supplemental Methods) before in-person confirmation of consent and specimen collection at the University of Maryland School of Public Health. For contacts, we collected a blood specimen at the first visit and measured oral temperature, blood oxygen saturation (SpO_2_), and collected a mid-turbinate swab (MTS) from each nostril and a saliva sample, at approximately two-day intervals.

Cohort members with a positive saliva sample, recently-diagnosed cases, and contacts with a positive test during follow-up were invited for viral shedding assessment visits on two days separated by one day. Venous blood was collected at the first visit; MTS, saliva, phone/tablet swab, and 30-minute G-II exhaled breath samples were collected at each visit[10,11]. To test the effectiveness of masks as source control, we asked participants to provide paired breath samples at each visit, first while wearing a mask and then without[13]. We provided a surgical mask at one visit and, to test masks in general use by the public, we tested the mask brought by the participant at the other; order of mask type was randomized to avoid bias. Cases studied before September 2020 were asked to repeat the alphabet three times within the 30-minute sampling period, as previously described [13]. Subsequent cases were asked to shout “Go Terps” 30 times and sing “Happy Birthday” loudly three times at 5, 15, and 25 minutes into each 30-minute sampling period. Participants with more severe symptoms sometimes opted to give only one 30-minute breath sample; for these participants, sampling without a face mask was given priority.

### Symptom evaluation

Symptoms were self-reported and measured on a scale of 0-3 and composite scores were sums of individual symptom scores for systemic, gastrointestinal, lower respiratory, and upper respiratory symptoms as previously described[10], see Supplementary Information [SI] for additional details.

### Sample preparation

MTS and phone/tablet swabs were eluted in 1 mL of Universal Transport Media (BD). MTS from both nostrils were combined. G-II coarse and fine aerosol samples were concentrated, resulting in a 1-mL final sample volume as previously described.[10] One aliquot each of MTS and saliva samples were kept at 4 °C for immediate qRT-PCR. All other samples were stored at −80 °C until further analysis.

### Laboratory analyses

Specific laboratory analysis details can be found in the SI. Cohort saliva was processed using the SalivaDirect method[14]. For all other samples, nucleic acids were extracted with MagMax Pathogen RNA/DNA Kit (Applied Biosystems) on KingFisher Duo Prime (Thermo Scientific), following manufacturer protocols specific to sample type. Viral RNA was detected and quantified using the TaqPath COVID-19 Real Time PCR Assay Multiplex and coinfection of samples was determined using the TaqMan Array Card (both Thermo Scientific). RNA copy numbers are reported per mL for saliva and per sample for all other sample types. The limit of detection (LOD) was 75 copies/sample and the limit of quantification (LOQ) was 250 copies/sample (SI).

Viral infectivity was measured by first propagating virus on Vero E6 cells stably expressing TMPRSS2[15] then transferring the media to A549 cells stably expressing human ACE2 (A549-ACE2, from BEI NR-53522 and a gift from Dr. Adolfo Garcia Sastre). Infected A549-ACE2 cells were quantified using immunofluorescence staining with anti-SARS-CoV-2 nucleocapsid antibody (Sino Biological 40143-R004) and Hoechst 33342 (Invitrogen H3570), and imaging with a Celigo Imaging Cytometer (Nexcelom) (Figures S1, S2). Plasma was analyzed for IgG, IgM, and IgA antibodies to SARS-CoV-2 using a modified protocol described by Stadlbauer et al., 2020[16]. MTS and saliva samples were sequenced at the Walter Reed Army Institute for Research or the University of Maryland Institute for Genome Sciences. SARS-CoV-2 lineages and mutations were determined using PANGOLIN and Nextstrain tools[17,18].

### Statistical analysis

We included analysis of all actively infected cases recruited from the campus and surrounding community over the course of one year. To ensure comparability of with and without mask samples in the analysis of mask efficacy, we included only paired same-day samples collected with and without a face mask from the same person and controlled for numbers of coughs during each sampling session.

Data curation, cleaning, analysis, and visualization were performed using R Studio and R (version 4.1.0)[19], with R packages lme4, lmec, and ggplot2[20]. We made group comparison between seronegative (no detectable antibody) and seropositive cases using two-sample t-test for continuous variables and Fisher’s exact test for categorical variables. To handle censored observations below the limit of detection, we applied linear mixed-effect models with censored responses[21,22] and included nested random effects of subject and sample nested within subjects. The geometric means and standard deviations of viral RNA copy numbers for all sample types and the effect of the predictors of viral RNA shedding were estimated from the model. Deidentified data and code for the accepted manuscript will be made available on github.

## Results

Sixty-one people with active SARS-CoV-2 infection (recent onset and positive PCR for MTS or saliva) were enrolled in breath testing: 3 from a weekly saliva testing cohort, 43 from 55 recently diagnosed COVID-19 cases, and 15 from among 62 contacts of cases followed to detect early onset of infection (Figure S3). Eight people (13%) with active infection had either IgM or IgG antibodies to SARS-CoV-2 spike protein at the first breath testing visit and four were unable to give a venous blood sample.

The 57 participants with known serologic status at the time of breath sampling (49 seronegative, 8 seropositive) were enrolled from zero to 12 days post onset of symptoms or first positive test (Table 1, Figure S4). All cases were asymptomatic or mild at the time of study. The seronegative cases included all participants with singing experience and alpha-variant infection – there were no other significant differences based on serologic status. No volunteer was taking antiviral medication at the time of breath sample collection and no co-infections were identified.

**Table 1.**
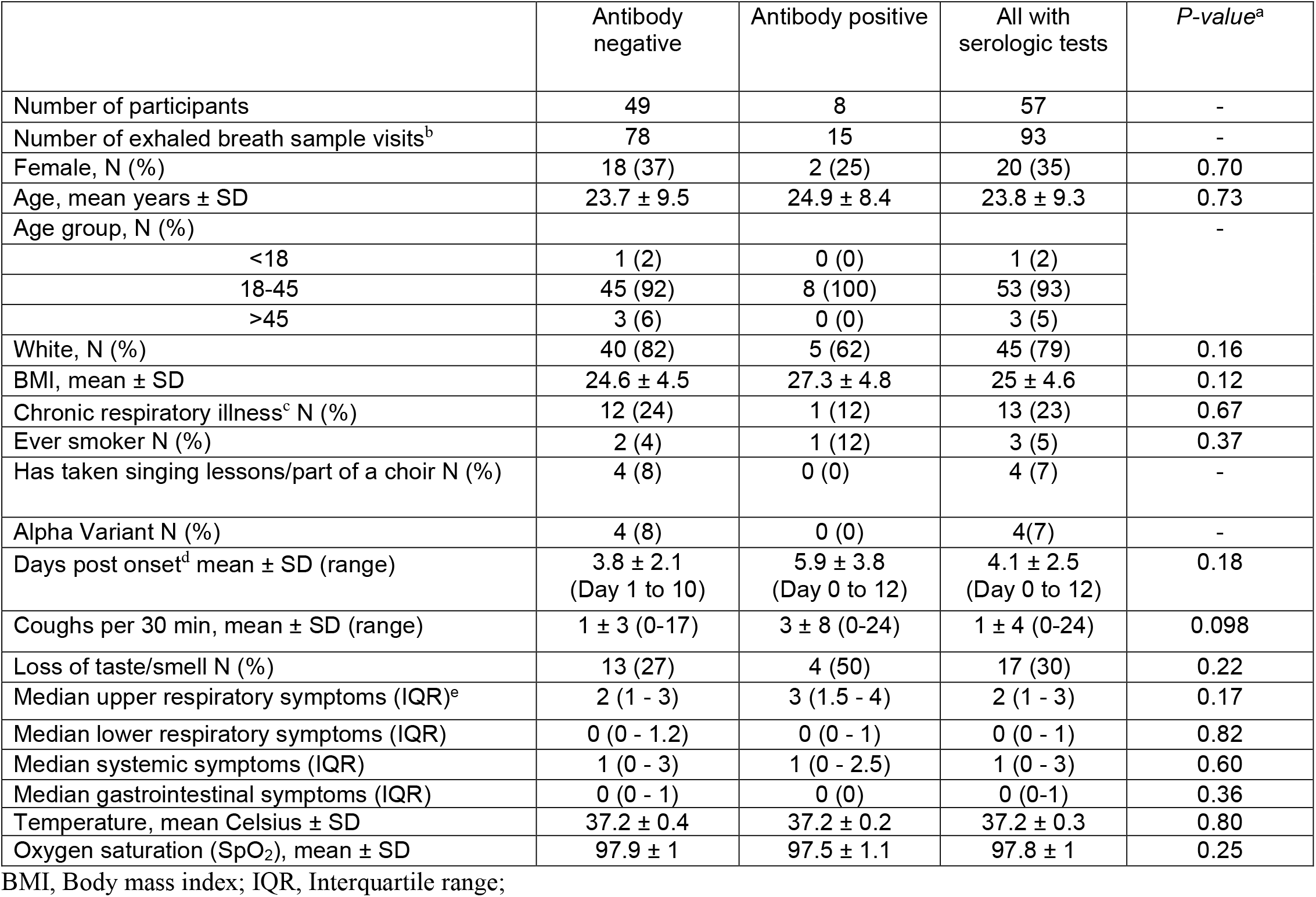
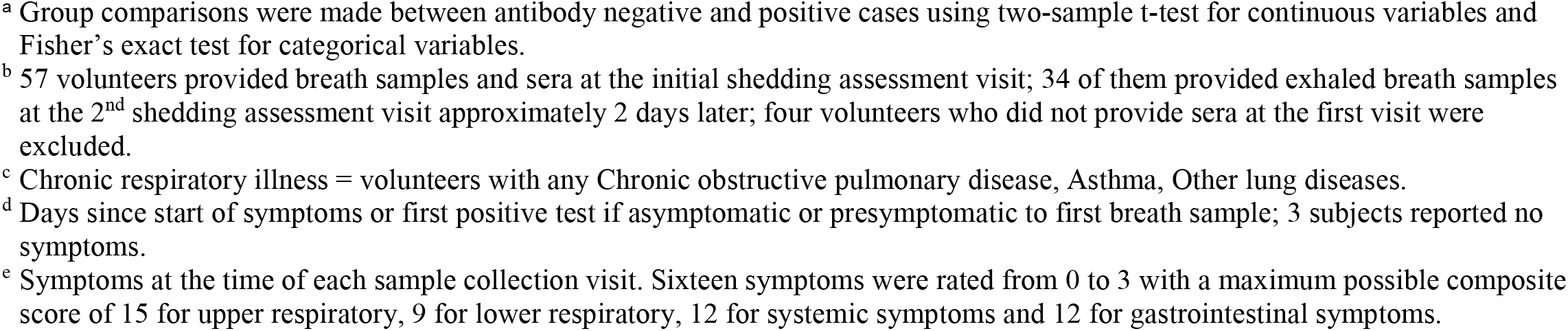
Characteristics of the study population.

|  | Antibody negative | Antibody positive | All with serologic tests | *P-value*<sup>a</sup> |
| --- | --- | --- | --- | --- |
| Number of participants | 49 | 8 | 57 | - |
| Number of exhaled breath sample visits<sup>b</sup> | 78 | 15 | 93 | - |
| Female, N (%) | 18 (37) | 2 (25) | 20 (35) | 0.70 |
| Age, mean years ± SD | 23.7 ± 9.5 | 24.9 ± 8.4 | 23.8 ± 9.3 | 0.73 |
| Age group, N (%) |  |  |  | - |
| <18 | 1 (2) | 0 (0) | 1 (2) |  |
| 18-45 | 45 (92) | 8 (100) | 53 (93) |  |
| >45 | 3 (6) | 0 (0) | 3 (5) |  |
| White, N (%) | 40 (82) | 5 (62) | 45 (79) | 0.16 |
| BMI, mean ± SD | 24.6 ± 4.5 | 27.3 ± 4.8 | 25 ± 4.6 | 0.12 |
| Chronic respiratory illness<sup>c</sup> N (%) | 12 (24) | 1 (12) | 13 (23) | 0.67 |
| Ever smoker N (%) | 2 (4) | 1 (12) | 3 (5) | 0.37 |
| Has taken singing lessons/part of a choir N (%) | 4 (8) | 0 (0) | 4 (7) | - |
| Alpha Variant N (%) | 4 (8) | 0 (0) | 4(7) | - |
| Days post onset<sup>d</sup> mean ± SD (range) | 3.8 ± 2.1<br>(Day 1 to 10) | 5.9 ± 3.8<br>(Day 0 to 12) | 4.1 ± 2.5<br>(Day 0 to 12) | 0.18 |
| Coughs per 30 min, mean ± SD (range) | 1 ± 3 (0-17) | 3 ± 8 (0-24) | 1 ± 4 (0-24) | 0.098 |
| Loss of taste/smell N (%) | 13 (27) | 4 (50) | 17 (30) | 0.22 |
| Median upper respiratory symptoms (IQR)<sup>e</sup> | 2 (1 - 3) | 3 (1.5 - 4) | 2 (1 - 3) | 0.17 |
| Median lower respiratory symptoms (IQR) | 0 (0 - 1.2) | 0 (0 - 1) | 0 (0 - 1) | 0.82 |
| Median systemic symptoms (IQR) | 1 (0 - 3) | 1 (0 - 2.5) | 1 (0 - 3) | 0.60 |
| Median gastrointestinal symptoms (IQR) | 0 (0 - 1) | 0 (0) | 0 (0-1) | 0.36 |
| Temperature, mean Celsius ± SD | 37.2 ± 0.4 | 37.2 ± 0.2 | 37.2 ± 0.3 | 0.80 |
| Oxygen saturation (SpO<sub>2</sub>), mean ± SD | 97.9 ± 1 | 97.5 ± 1.1 | 97.8 ± 1 | 0.25 |

BMI, Body mass index; IQR, Interquartile range;
<sup>a</sup> Group comparisons were made between antibody negative and positive cases using two-sample t-test for continuous variables and Fisher's exact test for categorical variables. <sup>b</sup> 57 volunteers provided breath samples and sera at the initial shedding assessment visit; 34 of them provided exhaled breath samples at the 2<sup>nd</sup> shedding assessment visit approximately 2 days later; four volunteers who did not provide sera at the first visit were excluded. <sup>c</sup> Chronic respiratory illness = volunteers with any Chronic obstructive pulmonary disease, Asthma, Other lung diseases. <sup>d</sup> Days since start of symptoms or first positive test if asymptomatic or presymptomatic to first breath sample; 3 subjects reported no symptoms. <sup>e</sup> Symptoms at the time of each sample collection visit. Sixteen symptoms were rated from 0 to 3 with a maximum possible composite score of 15 for upper respiratory, 9 for lower respiratory, 12 for systemic symptoms and 12 for gastrointestinal symptoms.

### Seronegative cases

Among seronegative participants, 4 (8%) were infected with the alpha and none with delta or other variants associated with increased transmissibility[23] (Table S1). Symptoms tended to be more pronounced at the first shedding visit, two participants were febrile at the second shedding visit (Figures S5-S6), and three were asymptomatic. Each of the 4 seronegative alpha variant cases had detectable concentrations of viral RNA in all MTS, saliva, fomite, and aerosol samples (Table 2). Among the remaining seronegative participants, we detected viral RNA in all MTS, 99% of saliva, 49% of fomite, 19% of coarse-, and 31% of fine-aerosol samples. The geometric mean concentrations of viral RNA were significantly greater for all sample types collected from alpha variant cases.

**Table 2.** Viral RNA from cases seronegative for SARS-CoV-2 antibodies at the first assessment

| Sample Type | Variant | Case<sup>a</sup> | Cases with $\geq 1$ positive sample<sup>b</sup><br>N (%) | | Samples | Positive Samples<sup>c</sup><br>N (%) | | GM (95% CI)<sup>d</sup> | Maximum<br>RNA<br>copies<sup>e</sup> |
| --- | --- | --- | --- | --- | --- | --- | --- | --- | --- |
| | | | $\geq \text{LOD}$ | $\geq \text{LOQ}$ | | $\geq \text{LOD}$ | $\geq \text{LOQ}$ | | |
| Mid-turbinate<br>swab | Alpha | 4 | 4 (100) | 4 (100) | 6 | 6 (100) | 6 (100) | $3.8 \times 10^8$<br>($3.3 \times 10^8$, $4.4 \times 10^8$) | $2.9 \times 10^9$ |
| | Other | 45 | 45 (100) | 45 (100) | 74 | 74 (100) | 73 (99) | $3.8 \times 10^6$<br>($1.4 \times 10^6$, $1.0 \times 10^7$) | $5.1 \times 10^9$ |
| Saliva | Alpha | 4 | 4 (100) | 4 (100) | 6 | 6 (100) | 6 (100) | $1.9 \times 10^7$<br>($2.7 \times 10^6$, $1.3 \times 10^8$) | $5.2 \times 10^8$ |
| | Other | 45 | 44 (98) | 44 (98) | 74 | 73 (99) | 70 (95) | $2.1 \times 10^5$<br>($8.1 \times 10^4$, $5.4 \times 10^5$) | $3.9 \times 10^8$ |
| Fomite | Alpha | 4 | 4 (100) | 4 (100) | 6 | 6 (100) | 4 (67) | 560 (530, 590) | $1.7 \times 10^4$ |
| | Other | 45 | 28 (62) | 14 (31) | 74 | 36 (49) | 17 (23) | 46 (17, 120) | $1.2 \times 10^6$ |
| Coarse<br>($>5 \mu\text{m}$)<br>Aerosol | Alpha | 4 | 4 (100) | 2 (50) | 6 | 6 (100) | 3 (50) | 140 (28, 730) | $5.1 \times 10^4$ |
| | Other | 45 | 11 (24) | 5 (11) | 72 | 14 (19) | 5 (7) | 7 (3.2, 15) | $2.6 \times 10^4$ |
| Fine<br>($\leq 5 \mu\text{m}$)<br>Aerosol | Alpha | 4 | 4 (100) | 3 (75) | 6 | 6 (100) | 4 (67) | 580 (450, 740) | $5.4 \times 10^4$ |
| | Other | 45 | 18 (40) | 8 (18) | 72 | 22 (31) | 9 (12) | 18 (9.1, 34) | $2.8 \times 10^3$ |

<sup>a</sup> Participants with a mid-turbinate or saliva samples positive for SARS-CoV-2 viral RNA by qRT-PCR and seronegative for SARS-CoV-2 spike protein antibody at enrollment and who provided at least one 30-minute sample of exhaled breath.
<sup>b</sup> Number of participants with at least one sample ≥ limit of detection (LOD) or ≥ limit of quantification (LOQ) (Supplementary.
<sup>c</sup> Samples positive and $\geq \text{LOD}$ had at least one replicate with confirmed amplification after inspection and quality control (LOD $\sim 75$ RNA copies with 95% probability of detection) and $\geq \text{LOQ}$ if the mean of replicate assays was $\geq 250$ RNA copies.
<sup>d</sup> GM = geometric mean. The GMs were computed, accounting for samples below the LOD, using a linear mixed-effects model for censored responses (R Project LMEC package) using data for all samples of each sample type with nested random effects of samples within study participant.
<sup>e</sup> The largest quantity of RNA copies detected based on the mean of replicates qRT-PCR aliquots.

### Exhaled breath viral RNA from seronegative cases without masks

Viral RNA content of 30-minute breath aerosol samples was similar to the amount of RNA recovered from participants’ mobile phones (Table 2). The frequency of detection of viral RNA in aerosols was greatest 2-5 days post onset of symptoms or first positive test (Figure S7) but was not a significant predictor of the quantity of viral RNA in the aerosols (Table 3). Viral RNA in MTS and saliva were moderately correlated (Figure S8: *ρ*=0.46) but only RNA in MTS was a strong predictor of viral load in aerosols (Table 3, Figures S9, S10). The quantity of viral RNA in the fine-aerosol fraction was 1.9-fold (95% confidence interval [CI] 1.2 to 2.9-fold) greater than in the coarse-aerosol fraction (not shown).

**Table 3.**
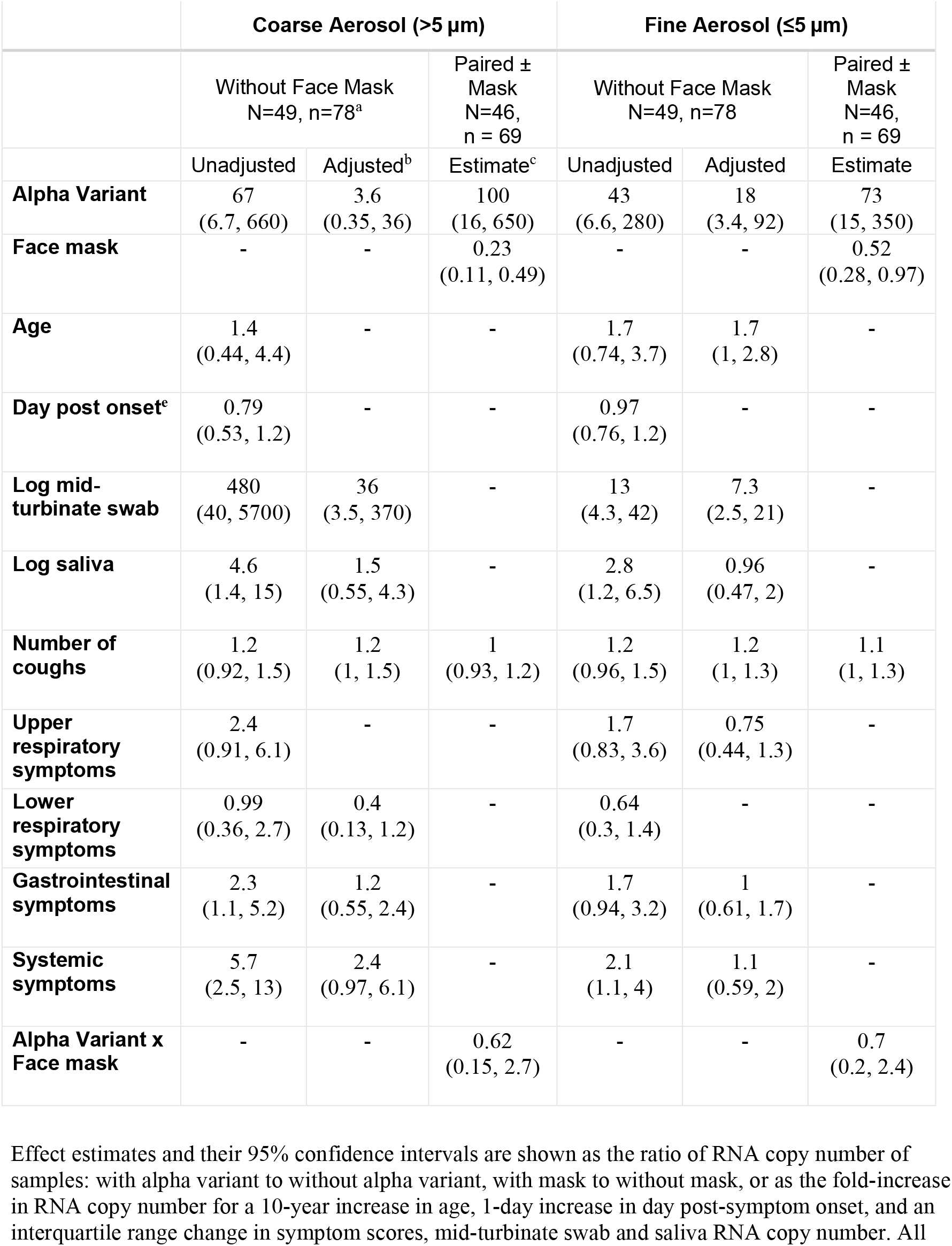
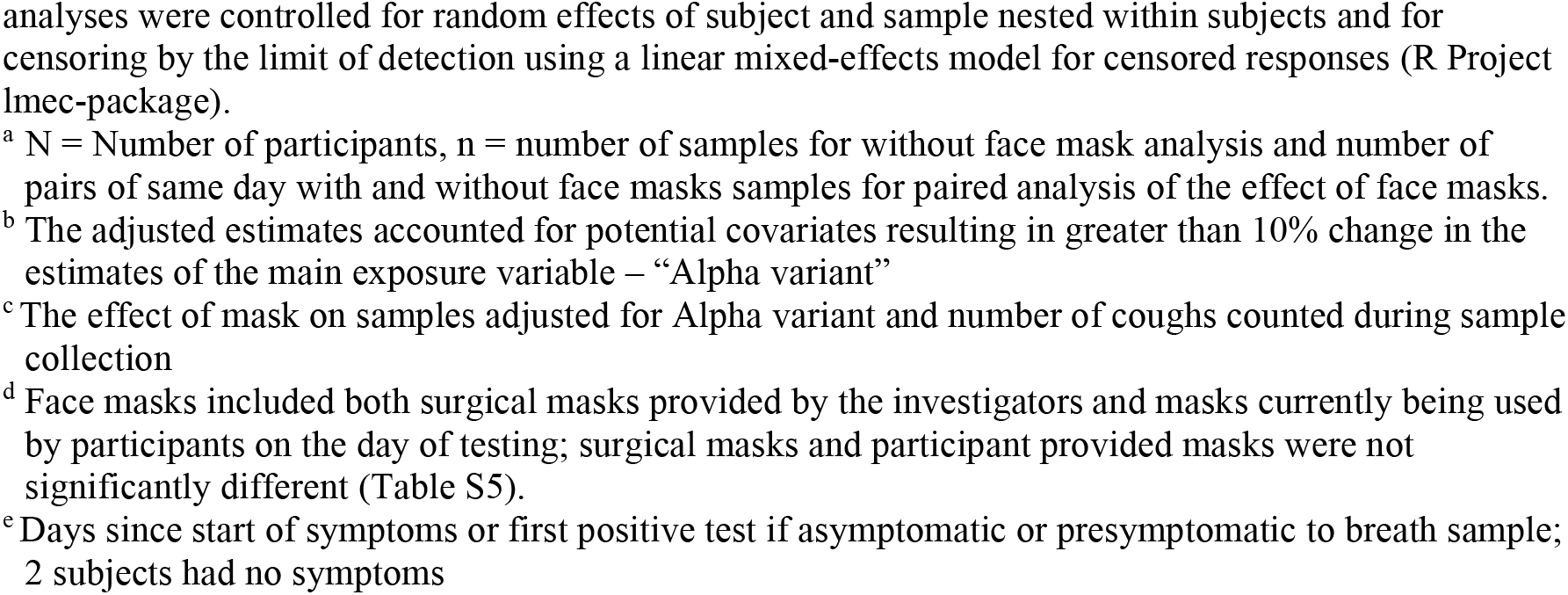
Predictors of viral RNA shedding among seronegative participants

|  | **Coarse Aerosol (>5 µm)** |  |  | **Fine Aerosol (≤5 µm)** |  |  |
| --- | --- | --- | --- | --- | --- | --- |
|  | Without Face Mask<br>N=49, n=78<sup>a</sup> |  | Paired ±<br>Mask<br>N=46,<br>n = 69 | Without Face Mask<br>N=49, n=78 |  | Paired ±<br>Mask<br>N=46,<br>n = 69 |
|  | Unadjusted | Adjusted<sup>b</sup> | Estimate<sup>c</sup> | Unadjusted | Adjusted | Estimate |
| **Alpha Variant** | 67<br>(6.7, 660) | 3.6<br>(0.35, 36) | 100<br>(16, 650) | 43<br>(6.6, 280) | 18<br>(3.4, 92) | 73<br>(15, 350) |
| **Face mask** | - | - | 0.23<br>(0.11, 0.49) | - | - | 0.52<br>(0.28, 0.97) |
| **Age** | 1.4<br>(0.44, 4.4) | - | - | 1.7<br>(0.74, 3.7) | 1.7<br>(1, 2.8) | - |
| **Day post onset<sup>e</sup>** | 0.79<br>(0.53, 1.2) | - | - | 0.97<br>(0.76, 1.2) | - | - |
| **Log mid-turbinate swab** | 480<br>(40, 5700) | 36<br>(3.5, 370) | - | 13<br>(4.3, 42) | 7.3<br>(2.5, 21) | - |
| **Log saliva** | 4.6<br>(1.4, 15) | 1.5<br>(0.55, 4.3) | - | 2.8<br>(1.2, 6.5) | 0.96<br>(0.47, 2) | - |
| **Number of coughs** | 1.2<br>(0.92, 1.5) | 1.2<br>(1, 1.5) | 1<br>(0.93, 1.2) | 1.2<br>(0.96, 1.5) | 1.2<br>(1, 1.3) | 1.1<br>(1, 1.3) |
| **Upper respiratory symptoms** | 2.4<br>(0.91, 6.1) | - | - | 1.7<br>(0.83, 3.6) | 0.75<br>(0.44, 1.3) | - |
| **Lower respiratory symptoms** | 0.99<br>(0.36, 2.7) | 0.4<br>(0.13, 1.2) | - | 0.64<br>(0.3, 1.4) | - | - |
| **Gastrointestinal symptoms** | 2.3<br>(1.1, 5.2) | 1.2<br>(0.55, 2.4) | - | 1.7<br>(0.94, 3.2) | 1<br>(0.61, 1.7) | - |
| **Systemic symptoms** | 5.7<br>(2.5, 13) | 2.4<br>(0.97, 6.1) | - | 2.1<br>(1.1, 4) | 1.1<br>(0.59, 2) | - |
| **Alpha Variant x Face mask** | - | - | 0.62<br>(0.15, 2.7) | - | - | 0.7<br>(0.2, 2.4) |

Effect estimates and their 95% confidence intervals are shown as the ratio of RNA copy number of samples: with alpha variant to without alpha variant, with mask to without mask, or as the fold-increase in RNA copy number for a 10-year increase in age, 1-day increase in day post-symptom onset, and an interquartile range change in symptom scores, mid-turbinate swab and saliva RNA copy number. All
analyses were controlled for random effects of subject and sample nested within subjects and for censoring by the limit of detection using a linear mixed-effects model for censored responses (R Project lme4-package).
<sup>a</sup> N = Number of participants, n = number of samples for without face mask analysis and number of pairs of same day with and without face masks samples for paired analysis of the effect of face masks.
<sup>b</sup> The adjusted estimates accounted for potential covariates resulting in greater than 10% change in the estimates of the main exposure variable – “Alpha variant”
<sup>c</sup> The effect of mask on samples adjusted for Alpha variant and number of coughs counted during sample collection
<sup>d</sup> Face masks included both surgical masks provided by the investigators and masks currently being used by participants on the day of testing; surgical masks and participant provided masks were not significantly different (Table S5).
<sup>e</sup> Days since start of symptoms or first positive test if asymptomatic or presymptomatic to breath sample; 2 subjects had no symptoms

### Effect of alpha variant infection on viral RNA shedding from seronegative cases

Alpha variant infection was associated with significantly greater amounts of viral RNA shedding than infection with earlier strains and variants. In bivariate analyses of samples collected without masks (Tables 3 and S3), the alpha variant was associated with significantly greater viral RNA shedding than wild-type and other variants not associated with increased transmissibility. Fine-aerosol shedding remained significantly greater for alpha variant infections (18-fold, 95% CI, 3.4 to 92-fold) after adjusting for the increased viral RNA in MTS and saliva, the number of coughs during sampling sessions, and symptoms (Table 3). We also analyzed the impact of alpha variants on shedding using the larger data set, including samples collected with and without masks. After controlling for the effect of masks and numbers of coughs during sampling, alpha variant infection was associated with a 100-fold (95% CI, 16 to 650-fold) increase in coarse- and a 73-fold (95% CI, 15 to 350-fold) increase in fine-aerosol RNA shedding (Table 3).

### Effect of masks on viral RNA shedding from seronegative cases

We observed statistically significant reductions in aerosol shedding regardless of mask type after adjusting for number of coughs during sampling sessions: 77% (95% CI, 51% to 89%) reduction for coarse and 48% (95% CI, 3% to 72%) for fine aerosols (Figure 1). Surgical masks did not perform significantly differently than others (Table S5), and mask performance was not significantly different for alpha variant infections (Table 3). The types of face masks brought by participants varied and progressed from single-layer homemade cloth masks to more substantial double-layer cloth masks, surgical masks, double masks, and a KN95 over the course of the year (Table S4).

**Figure 1.**
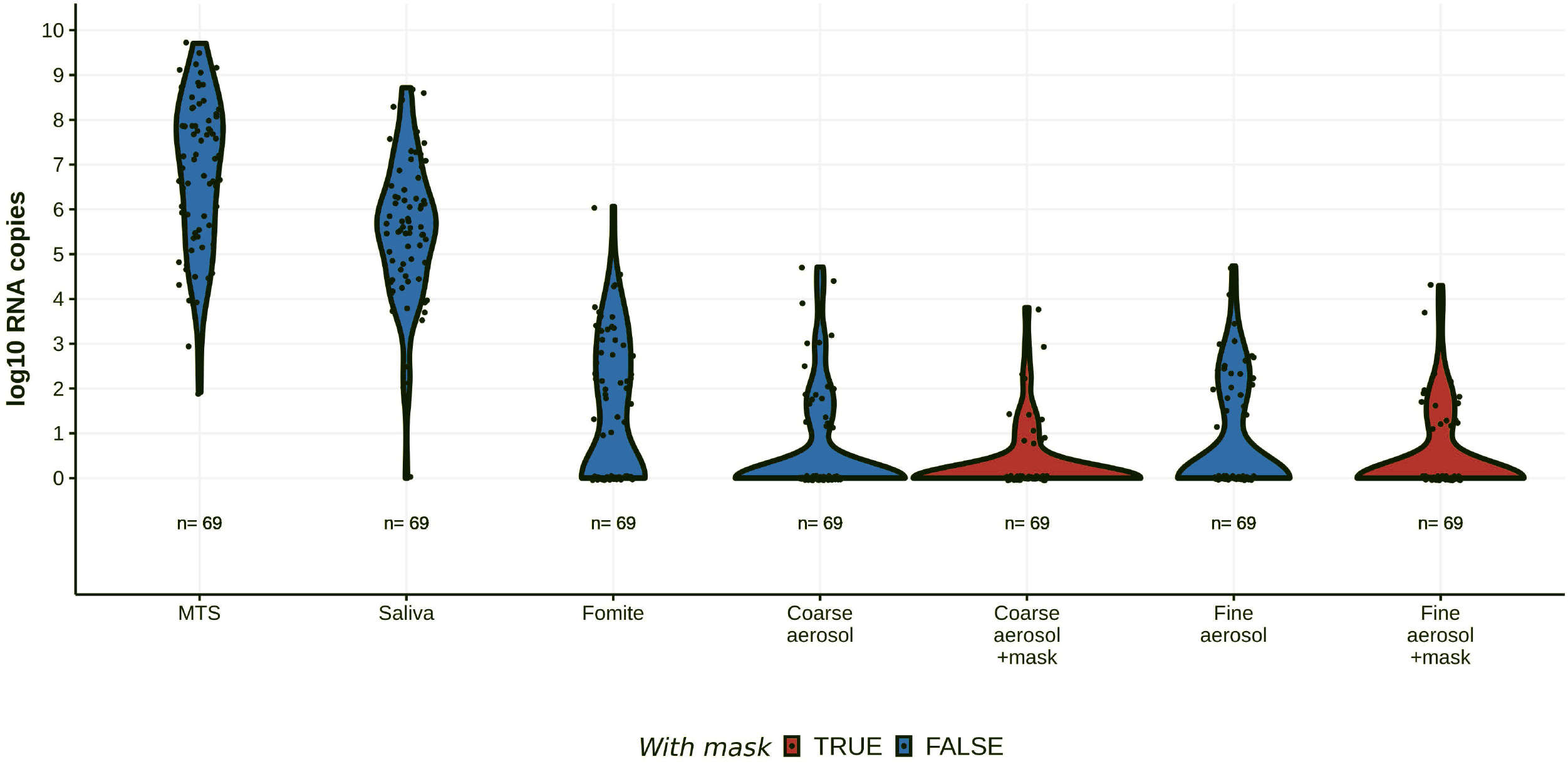
Viral RNA shedding in paired with and without face mask samples. Viral RNA measured during 69 same-day paired sampling events with and without mask from 46 seronegative cases. Samples with no detected viral RNA were assigned a copy number value of one. Exhaled breath aerosols were obtained in 30-minute sampling durations. “+mask” = sample collected while wearing a face mask. MTS = mid-turbinate swab, Fomite = swab of participant’s mobile phone.

### Seropositive cases

Eight participants had antibodies to SARS-CoV-2 spike protein at the time of breath sample collection. Seropositive cases tended to cough more than seronegative cases (Table 1) but none of the exhaled aerosol samples from seropositive cases had detectable viral RNA (Table S6).

### Virus cultures

Among samples subjected to virus culture, we detected infectious virus (Figure S2) in 50 (68%) of 73 MTS and 20 (32%) of 62 saliva samples from seronegative participants (Figure 2, Table S2a). The RNA concentration associated with a 50% probability of a positive culture was 7.8 ⨯ 10^5^ for MTS and 5.2 ⨯ 10^6^ for saliva (Figure S11, Table S7). None of the 75 fine-aerosol samples collected while not wearing face masks were culture-positive. Two (3%) of the 66 fine-aerosol samples collected from participants while wearing face masks were culture-positive, including one from a person infected with the alpha variant 2 days post onset and one from a person with a Nextstrain clade 20G virus 3 days post onset. Fomite and coarse-aerosol samples subjected to culture were negative.

**Figure 2.**
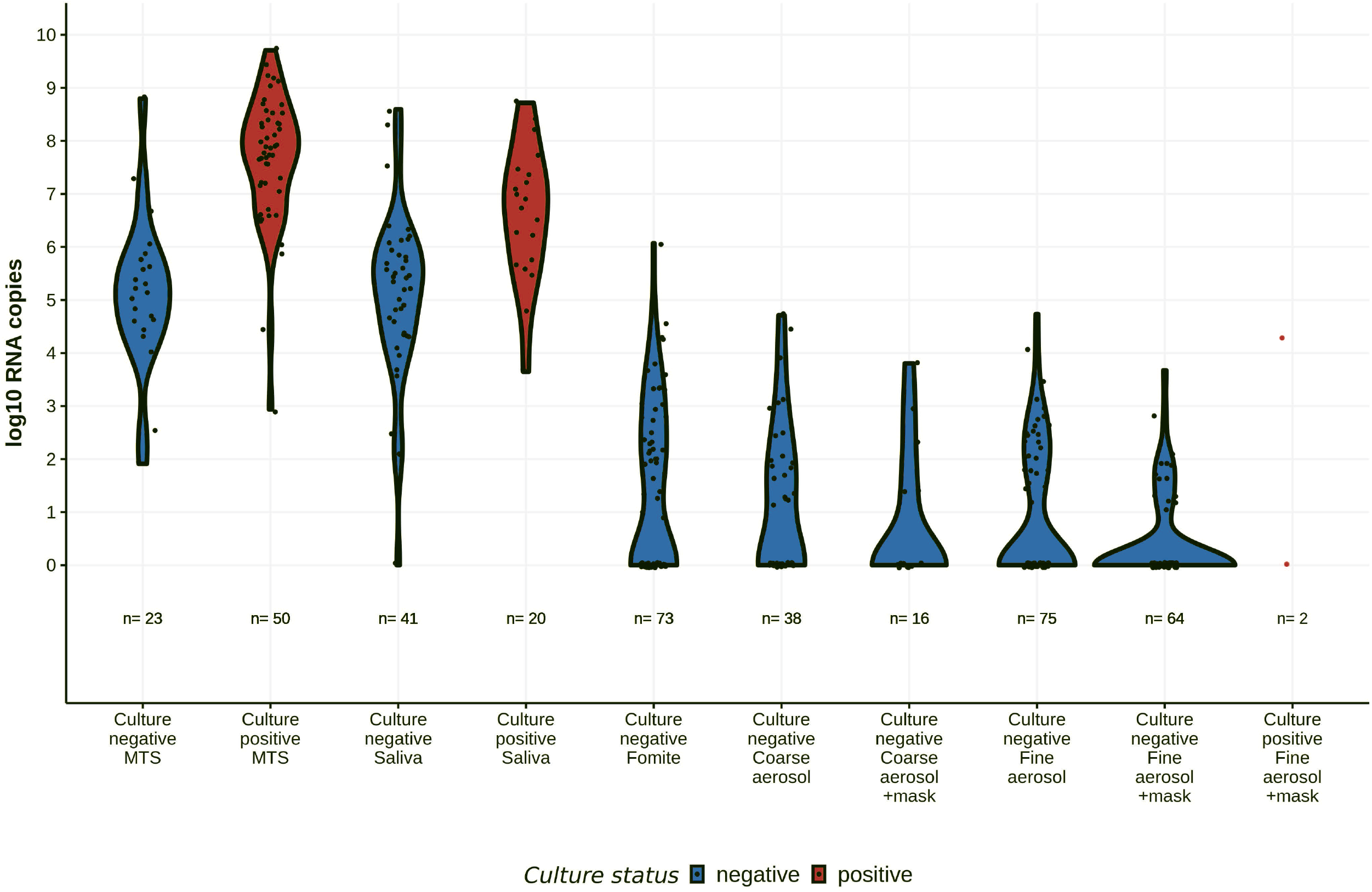
Viral RNA content and culture results of samples from all sampling events for seronegative cases. Samples with no detected viral RNA were assigned a copy number value of one. Exhaled breath aerosols were obtained during 30-minute sampling events and included unpaired with and without face mask samples. Five fine aerosol samples with face mask and three fomite samples were not available for culture. A subset of MTS, saliva, and coarse aerosol samples were subjected to culture. MTS = mid-turbinate swab, Fomite = swab of participant’s mobile phone.

## Discussion

Alpha variant infection yielded one to two orders of magnitude more viral RNA in exhaled breath when compared with earlier strains and variants not associated with increased transmissibility. Our observation of increased aerosol shedding, even after controlling for the increased amounts of viral RNA in the nose and mouth, suggests that evolutionary pressure is selecting for SARS-CoV-2 capable of more efficient aerosol generation.

We recovered infectious virus from two exhaled breath fine-aerosol samples, approximately two-thirds of MTS, and one-third of saliva samples. Logistic regression analysis of the MTS and saliva samples suggests that there is a small but measurable probability that each RNA copy represents an infectious viral particle consistent with previous dose-response models of infectious viruses[24]. The probability of detecting infectious virus in our cell culture system may be as high as 10^−4^ per RNA copy in saliva samples with 10^2^ copies. We recovered infectious virus from one of the two fine-aerosol samples with >10^4^ RNA copies per sample, where the estimated probability of a positive culture was 8% per sample, based on the regression model. The other culture-positive aerosol sample was one of 98 aerosol samples below the 75-copy limit of detection for RNA. This observation is consistent with a previous report of infectious SARS-CoV-2 in an aerosol sample with very low RNA copy numbers[5], and may suggest that virus particles in respiratory aerosols contain fewer defective virions than MTS, or that the fluid is less likely to interfere with viral cell attachment and entry than are nasal secretions and saliva. It is likely that human airway epithelium is more susceptible to infection than laboratory cell cultures[25].

Loose-fitting face masks, including surgical masks and those in daily use by the study participants, produced significant, albeit modest, reductions in the amount of viral RNA in exhaled breath, supporting their use as source control. Consistent with previous studies of influenza, SARS-CoV-2 viral RNA was shed more abundantly in fine than coarse aerosol and masks were more effective at blocking release of coarse aerosol[10,13,26,27]. The effectiveness of face masks for blocking release of fine aerosol (1.9-fold) was similar to the effectiveness we previously reported for influenza (2.8-fold). However, the 4.3-fold (or 77%) reduction in coarse aerosol was significantly less than the 25-fold reduction previously observed for influenza shedding[13]. This discrepancy likely arose because the more vigorous and extended vocalizations in the current study would be expected to maximize leakage around loose-fitting face masks. Face masks worked equally well for containing alpha variant and “wild-type” aerosol shedding.

Viral RNA aerosol collected from 22 COVID-19 patients in Singapore using a G-II measured similar RNA copy numbers and overall rates of positive breath samples (59% versus 51% here) during singing and loud talking as observed here[28]. A majority (68%) of the cases studied in Singapore were infected by variants associated with increased transmissibility, while 18% of infections were not variants of concern or interest, and each case was sampled on only one day. By comparison, our sample included more “wild-type” infections and sampling days per person allowing analysis of the impact of variants on shedding. One delta variant was studied in Singapore and none in this study. The shedding rates detected using the G-II in both studies, however, were lower than those reported by Ma et al. using a sampler that requires subjects to blow through a narrow straw[29]. Ma et al. and our data are in agreement that among persons infected with “wild-type” strains, a minority (26% and 31%, respectively) shed detectable levels of viral RNA into aerosols. However, our data on alpha variant infections and the data from Singapore suggest that this is changing and most cases may now be shedding viral aerosols more frequently. We have yet to enroll cases with the delta variant and study them using the Maryland protocol to test the expectation, based on our current findings, that delta will be associated with an additional large increase in aerosol shedding.

Our study had several limitations. Although we attempted to identify and test cases early through weekly testing of a cohort and intensive follow-up of close contacts, most cases were still studied several days after onset of symptoms or first positive test. This likely resulted in missing the most contagious period[30–33]. Furthermore, all cases were mild at the time of testing and only two went on to have moderate severity and require hospitalization. As a result, our data may not represent the upper end of the spectrum of shedding. The types of face masks worn by participants changed over the course of the study, as did the quality of surgical masks that we could purchase. Therefore, we cannot report on the efficacy of specific loose-fitting masks. This work does, however, provide information on the average amount of source control provided by community masking. Finally, logistical considerations required that we freeze samples between collection and culture, with potential loss of infectiousness[34], especially for dilute aerosol specimens.

Overall, our results demonstrate that people with mild or asymptomatic SARS-CoV-2 infections released infectious aerosols in their exhaled breath. Face masks provided significant source control suggesting that community-wide masking even with loose-fitting masks can reduce viral aerosols in indoor air by half, making a significant contribution to reducing the spread of COVID-19. Our data also suggest that the virus is evolving toward more effective dissemination through aerosols and demonstrate that infectious virus can escape from loose-fitting masks. With the dominance of newer, more contagious variants than those we studied, increased attention to improved ventilation, filtration, air sanitation, and use of high-quality tight-fitting face masks or respirators (e.g., tight-fitting ASTM F3502-21 level-2 face-coverings or respirators) for respiratory protection will be increasingly important for controlling the pandemic. This will be especially true where vaccination rates are low, vaccine is not available, and for people with poor immune responses or waning immunity. Therefore, our data support community mask mandates and tight-fitting masks or respirators for workers in healthcare but also in all workplaces where people are sharing indoor air or have frequent public contact.

## Data Availability

Deidentified data and code for the accepted manuscript will be made available on github.

## Acknowledgements

We thank Dr. Jacques Ravel, Dr. Luke Tallon, and the Institute of Genome Sciences at the University of Maryland School of Medicine for performing deep sequencing of SARS-CoV-2 samples. We also thank Dr. Jamal Fadul and his clinic in College Park, Maryland, for assistance in recruiting study participants.

## Disclaimers and Funding

Material has been reviewed by the Walter Reed Army Institute of Research. There is no objection to its presentation and/or publication. The investigators have adhered to the policies for protection of human subjects as prescribed in AR 70–25. The View(s) expressed are those of the authors and do not necessarily reflect the official views of the Department of Health and Human Services, the Departments of the Army, Navy or Air Force, the Department of Defense, or the U.S. Government. The findings and conclusions in this report are those of the authors and do not necessarily represent the official position or policy of these agencies and no official endorsement should be inferred.

This work was supported by Prometheus-UMD, sponsored by the Defense Advanced Research Projects Agency (DARPA) BTO under the auspices of Col. Matthew Hepburn through agreement N66001-18-2-4015. This work was also supported by the National Institute of Allergy and Infectious Diseases Centers of Excellence for Influenza Research and Surveillance (CEIRS), Grant Number HHSN272201400008C, the Centers for Disease Control and Prevention, Contract Number 200-2020-09528. The findings and conclusions in this report are those of the authors and do not necessarily represent the official position or policy of these funding agencies and no official endorsement should be inferred.

This work was also supported by a grant from the Bill & Melinda Gates Foundation, and a generous gift from The Flu Lab (https://theflulab.org). The funders had no role in study design, data collection, analysis, decision to publish, or preparation of the manuscript.

None of the authors have a potential conflicting interest or funding source.

## Notes

### Competing Interest Statement

The authors have declared no competing interest.

### Clinical Trial

NA

### Author Declarations

This study was approved by the University of Maryland Institutional Review Board and the Human Research Protection Office of the Department of the Navy.

### Summary of Updates

Minor revisions and clarifications and some editing for brevity.

## References

1. World Health Organization. Coronavirus disease (COVID-19): How is it transmitted? 2021. Available at: https://www.who.int/news-room/q-a-detail/coronavirus-disease-covid-19-how-is-it-transmitted. Accessed 31 May 2021.

2. CDC. Scientific Brief: SARS-CoV-2 Transmission. 2021. Available at: https://www.cdc.gov/coronavirus/2019-ncov/science/science-briefs/sars-cov-2-transmission.html. Accessed 8 May 2021.

3. Morawska L, Milton DK. It Is Time to Address Airborne Transmission of Coronavirus Disease 2019 (COVID-19). Clin Infect Dis 2020; 71:2311–2313.

4. Greenhalgh T, Jimenez JL, Prather KA, Tufekci Z, Fisman D, Schooley R. Ten scientific reasons in support of airborne transmission of SARS-CoV-2. Lancet 2021; 397:1603–1605.

5. Lednicky JA, Lauzardo M, Fan ZH, et al. Viable SARS-CoV-2 in the air of a hospital room with COVID-19 patients. Int J Infect Dis 2020; 100:476–482.

6. Lednicky JA, Lauzardo M, Alam MdM, et al. Isolation of SARS-CoV-2 from the air in a car driven by a COVID patient with mild illness. Infectious Diseases (except HIV/AIDS), 2021. Available at: http://medrxiv.org/lookup/doi/10.1101/2021.01.12.21249603. Accessed 2 February 2021.

7. Chia PY, Coleman KK, Tan YK, et al. Detection of air and surface contamination by SARS-CoV-2 in hospital rooms of infected patients. Nat Commun 2020; 11:2800.

8. Liu Y, Ning Z, Chen Y, et al. Aerodynamic analysis of SARS-CoV-2 in two Wuhan hospitals. Nature 2020; 582:557–560.

9. Miller SL, Nazaroff WW, Jimenez JL, et al. Transmission of SARS-CoV-2 by inhalation of respiratory aerosol in the Skagit Valley Chorale superspreading event. Indoor Air 2020;

10. Yan J, Grantham M, Pantelic J, et al. Infectious virus in exhaled breath of symptomatic seasonal influenza cases from a college community. Proc Natl Acad Sci USA 2018; 115:1081–1086.

11. McDevitt JJ, Koutrakis P, Ferguson ST, et al. Development and Performance Evaluation of an Exhaled-Breath Bioaerosol Collector for Influenza Virus. Aerosol Sci Technol 2013; 47:444–451.

12. Harris PA, Taylor R, Minor BL, et al. The REDCap consortium: Building an international community of software platform partners. Journal of Biomedical Informatics 2019; 95:103208.

13. Milton DK, Fabian MP, Cowling BJ, Grantham ML, McDevitt JJ. Influenza virus aerosols in human exhaled breath: particle size, culturability, and effect of surgical masks. PLoS Pathog 2013; 9:e1003205.

14. Vogels CBF, Brackney D, Wang J, et al. SalivaDirect: Simple and sensitive molecular diagnostic test for SARS-CoV-2 surveillance. Infectious Diseases (except HIV/AIDS), 2020. Available at: http://medrxiv.org/lookup/doi/10.1101/2020.08.03.20167791. Accessed 17 August 2020.

15. Matsuyama S, Nao N, Shirato K, et al. Enhanced isolation of SARS-CoV-2 by TMPRSS2-expressing cells. PNAS 2020; 117:7001–7003.

16. Stadlbauer D, Amanat F, Chromikova V, et al. SARS-CoV-2 Seroconversion in Humans: A Detailed Protocol for a Serological Assay, Antigen Production, and Test Setup. Current Protocols in Microbiology 2020; 57:e100.

17. O’Toole Á, Hill V, McCrone JT, Scher E, Rambaut A. cov-lineages/pangolin. CoV-lineages, 2021. Available at: https://github.com/cov-lineages/pangolin. Accessed 15 July 2021.

18. Trevor Bedford, Richard Nehe, James Hadfield, et al. Nextstrain SARS-CoV-2 resources. 2020. Available at: https://nextstrain.org/sars-cov-2/. Accessed 29 July 2021.

19. R Core Team. R: A Language and Environment for Statistical Computing. Vienna, Austria: R Foundation for Statistical Computing, 2021. Available at: https://www.R-project.org/.

20. Wickham H. ggplot2: Elegant Graphics for Data Analysis. 2nd ed. Springer International Publishing, 2016. Available at: https://www.springer.com/gp/book/9783319242750. Accessed 11 June 2020.

21. Vaida F, Fitzgerald AP, DeGruttola V. Efficient Hybrid EM for Linear and Nonlinear Mixed Effects Models with Censored Response. Comput Stat Data Anal 2007; 51:5718–5730.

22. Vaida F, Liu L. Fast Implementation for Normal Mixed Effects Models With Censored Response. J Comput Graph Stat 2009; 18:797–817.

23. CDC. SARS-CoV-2 Variant Classifications and Definitions. 2021. Available at: https://www.cdc.gov/coronavirus/2019-ncov/variants/variant-info.html. Accessed 12 July 2021.

24. Haas CN. Estimation of risk due to low doses of microorganisms: a comparison of alternative methodologies. Am J Epidemiol 1983; 118:573–582.

25. Pohl MO, Busnadiego I, Kufner V, et al. SARS-CoV-2 variants reveal features critical for replication in primary human cells. PLoS Biol 2021; 19:e3001006.

26. Lindsley WG, Noti JD, Blachere FM, et al. Viable influenza A virus in airborne particles from human coughs. J Occup Environ Hyg 2015; 12:107–113.

27. Leung NHL, Chu DKW, Shiu EYC, et al. Respiratory virus shedding in exhaled breath and efficacy of face masks. Nat Med 2020; 26:676–680.

28. Coleman KK, Tay DJW, Tan KS, et al. Viral Load of SARS-CoV-2 in Respiratory Aerosols Emitted by COVID-19 Patients while Breathing, Talking, and Singing. medRxiv 2021; :2021.07.15.21260561.

29. Ma J, Qi X, Chen H, et al. COVID-19 patients in earlier stages exhaled millions of SARS-CoV-2 per hour. Clin Infect Dis 2020; Available at: https://academic.oup.com/cid/advance-article/doi/10.1093/cid/ciaa1283/5898624.

30. He X, Lau EHY, Wu P, et al. Temporal dynamics in viral shedding and transmissibility of COVID-19. Nature Medicine 2020; 26:672–675.

31. Yang Q, Saldi TK, Gonzales PK, et al. Just 2% of SARS-CoV-2-positive individuals carry 90% of the virus circulating in communities. Proc Natl Acad Sci U S A 2021; 118:e2104547118.

32. Jones TC, Biele G, Mühlemann B, et al. Estimating infectiousness throughout SARS-CoV-2 infection course. Science 2021; Available at: https://science.sciencemag.org/content/early/2021/05/24/science.abi5273. Accessed 26 May 2021.

33. Sun K, Wang W, Gao L, et al. Transmission heterogeneities, kinetics, and controllability of SARS-CoV-2. Science 2021; 371:eabe2424.

34. Huang C-G, Lee K-M, Hsiao M-J, et al. Culture-Based Virus Isolation To Evaluate Potential Infectivity of Clinical Specimens Tested for COVID-19. J Clin Microbiol 2020; 58:e01068–20.

