## Supplementary Information for "Infectious SARS-CoV-2 in Exhaled Aerosols and Efficacy of Masks During Early Mild Infection"

### Table of Contents

*Supplemental methods* 3

**Questionnaire** 3

**Symptom evaluation** 3

**Cohort Saliva Testing** 4

**Quantification of SARS-CoV-2 RNA** 4

**Detection of co-infection** 4

**Mammalian cell culture** 4

**SARS-CoV-2 viral culture** 5

**Immunofluorescence staining and microscopy** 5

**Antibody response assessment** 5

**SARS-CoV-2 sequencing and genome assembly** 6

**Additional statistical analysis methods** 6

*Supplemental Tables* 7

**Table S1. Variant Classification** 7

**Table S2A. Viral RNA copies and culture of samples from cases seronegative for SARS-CoV-2 antibodies at the first shedding assessment.** 8

**Table S2B. Viral RNA copies and culture of samples from alpha variant cases, seronegative for SARS-CoV-2 antibodies at the first assessment.** 9

**Table S2C. Viral RNA copies and culture of samples from wild type and other (non-alpha) variant cases, seronegative for SARS-CoV-2 antibodies at the first assessment.** 10

**Table S3. The effect of alpha variant on viral load of Mid-turbinate swab and Saliva** 11

**Table S4. Frequency of different mask types at the sample level used in the paired analyses** 12

**Table S5. Regression Coefficients for Effect of Masks and Mask Type** 12

**Table S6. Viral RNA copy numbers in participants who were already seropositive at enrollment.** 13

**Table S7. Probability of positive cultures at GM viral RNA copy numbers observed.** 15

*Supplemental Figures* 17

**Figure S1. Culture methods and sensitivity** 17

**Figure S2. Culture results from representative samples** 18

**Figure S3. Study design** 19

**Figure S4. Days since onset** 20

**Figure S5. Symptoms** 21

**Figure S6. Temperature** 22
**Figure S7. Positive and negative samples by day since onset.** 23
**Figure S8. Correlation between saliva and mid-turbinate swab RNA measurements** 24
**Figure S9. Correlation between the SARS-CoV-2 RNA copy numbers detected by qRT-PCR in upper respiratory samples (saliva and mid-turbinate swabs) and exhaled breath samples** 25
**Figure S10. Correlation between the cycle threshold (Ct) values for SARS-CoV-2 N-gene in the upper respiratory samples (saliva and mid-turbinate swabs) and exhaled breath samples.** 26
**Figure S11. Probability of a positive SARS-CoV-2 culture.** 27
***References*** *28*

**University of Maryland StopCOVID Research Group**
Donald K. Milton, MD, Dr.P.H.[a], Principal Investigator
Oluwasanmi Oladapo Adenaiye, M.B.B.S., M.S.[a], Barbara Albert, M.D., M.P.H. [a], P. Jacob Bueno de Mesquita, Ph.D. [a], Yi Esparza, B.S., R.N.[a], Jennifer German, Ph.D. [a], Filbert Hong, Ph.D. [a], Aaron Kassman, B.A.[a], Jianyu Lai, B.Med., M.P.H. [a], Michael Lutchenkov, B.S.[a], Kathleen McPhaul, R.N., M.P.H., Ph.D.[a], Dewansh Rastogi, M.Tech.[b], Maria Schanz, B.S.[a], Isabel Sierra Maldonado, M.H.A.[a], Aditya Srikakulapu, M.S.[a], Delwin Suraj, B.S.[a], S.-H. Sheldon Tai, Ph.D. [a], Faith Touré, B.S.[a], Rhonda Washington-Lewis[a], Somayeh Youssefi, Ph.D. [a], Stuart Weston, Ph.D.[c], Matthew Frieman, Ph.D.[c], Mara Cai B.S.[d], Ashok Agrawala, Ph.D.[d]

[a] Public Health Aerobiology and Biomarker Laboratory, Institute for Applied Environmental Health, University of Maryland School of Public Health, College Park, Maryland
[b] Department of Chemical and Biomolecular Engineering, A. James Clark School of Engineering, University of Maryland, College Park, Maryland
[c] Department of Microbiology and Immunology, University of Maryland School of Medicine, Baltimore, Maryland
[d] Department of Computer Science, University of Maryland, College Park, Maryland

### Supplemental methods

#### Questionnaire

Participants provided baseline information about demographics, medical history, tobacco and alcohol use, stress level, sleep and physical activity, singing experience, residential conditions, frequency of recent essential activities outside the home, and a standardized symptom checklist for symptoms over the previous seven days via an online survey at the time of study enrollment. Subjects who enrolled directly as cases provided this information just prior to their first study visit, whereas contacts or participants in the weekly testing cohort who later tested positive for SARS-CoV-2 may have provided it several days to weeks in advance of their first viral shedding visit. All participants answered an online questionnaire to update their current symptoms and medications, close contacts within the preceding 48 hours, essential activities with potential exposures outside the home in the last 48 hours, and time spent in their rooms and bedrooms just prior to or during each viral shedding study visit.

#### Symptom evaluation

Symptoms were self-reported and measured on a scale of 0-3 (0 = “no symptoms,” 1 = “just noticeable,” 2 = “clearly bothersome from time to time, but didn’t stop me from participating in activities,” 3 = “quite bothersome most or all of the time, and stopped me from participating in activities”). Composite scores were calculated by summing individual symptom scores, in the following categories: Systemic (max score of 12) = malaise + headache + muscle/joint ache + sweats/fever/chills; Gastrointestinal (max score of 12) = loss of appetite + nausea + vomit + diarrhea; Lower Respiratory (max score of 9) = chest tightness + shortness of breath + cough; Upper Respiratory (max score of 15) = runny nose + stuffy nose + sneeze + earache + sore throat).

Cohort Saliva Testing

Saliva samples were processed using the SalivaDirect method[1]. Briefly, 50 µL of individual saliva samples were treated with Proteinase K (New England Biolabs) and heated at 95°C for 5 minutes. 10 µL of heat-treated sample was then combined into pools of 3. qRT-PCR was set up on the same day; each reaction consisted of 1X TaqPath 1-Step Master Mix, No ROX, 1X TaqPath COVID-19 Real Time PCR Assay Multiplex (both from Thermo Scientific), and 10 µL of pooled, heat-treated saliva. Pooled samples were considered positive if at least 2 of the 3 target genes amplified. Positive pools were de-convoluted to determine the positive individual sample with 10 µL of the heat-treated saliva using the same TaqPath COVID-19 Real Time PCR Assay.

Quantification of SARS-CoV-2 RNA

Nucleic acids were extracted with MagMax Pathogen RNA/DNA Kit (Applied Biosystems) on KingFisher Duo Prime (Thermo Scientific), following the manufacturers' protocols specific to sample type. Total nucleic acid from 200 µL of each mid-turbinate swab (MTS), phone swab, cone swab, and G-II aerosol sample and 240 µL of each saliva sample was eluted in 50 µL of Elution Buffer and kept at 4 °C. MS2 phage was spiked in each extraction to control for extraction and PCR failure. qRT-PCR was set up on the same day; each reaction consisted 1X TaqPath 1-Step Master Mix, No ROX, 1X TaqPath COVID-19 Real Time PCR Assay Multiplex (both from Thermo Scientific), and 10 µL of eluted nucleic acids. All reactions were carried out in duplicate. Each PCR plate contained a positive control provided by the Assay and a no template control. The Ct values of the N gene assay were standardized against serial dilutions of RNA, which were purified from SARS-CoV-2 culture and quantified against inactivated SARS-CoV-2 (BEI Resources NR-52286). MS2 amplification was confirmed in all reactions that contain SARS-CoV-2 negative samples. S-gene target failure (SGTF) was defined as absence of S-gene amplification in the presence of N- and ORF1ab-gene amplification from MTS and saliva.

Sample volumes of breath aerosols were adjusted to 1 mL, each MTS was extracted in 1 mL, and all results were reported as total RNA copies per sample, or per mL for saliva. The limit of detection (LOD), the quantity giving a ≥95% probability of detection, was estimated to be 3 genome equivalents per reaction, giving an LOD for of 62 copies/mL for saliva and 75 copies/sample for all other sample types. The limit of quantification (LOQ), the quantity giving a coefficient of variation <35%, was 10 genome equivalents per reaction, which translated to an LOQ of 208 copies/mL for saliva and 250 copies/sample for all the other sample types.

Detection of co-infection

Nucleic acids were extracted with MagMax Pathogen RNA/DNA Kit (Applied Biosystems) on KingFisher Duo Prime (Thermo Scientific), following the manufacturers' protocols specific to sample type. Total nucleic acid from 200 µL of each MTS was eluted in 50 µL of Elution Buffer and kept at 4 °C. qRT-PCR was set up on the same day using the TaqMan Array Card (ThermoFisher Scientific) and TaqMan Fast Virus One Step Master Mix (Applied Biosystems). The array consists of the manufacterer's proprietary primer-probes designed to probe for 46 common respiratory viruses and bacteria.

Mammalian cell culture

A549 cells stably expressing human ACE2 (A549-ACE2, from BEI NR-53522 and a gift from Dr. Adolfo Garcia Sastre) were cultured in DMEM (Corning), supplemented with 10% (v/v) fetal calf serum

(FCS, Sigma) and 1% (v/v) penicillin/streptomycin (pen/strep, 10,000 U/ml / 10 mg/ml; Gemini Bioscience). Vero E6 cells stably expressing TMPRSS2 (VeroTMPRSS2)[2] were cultured in DMEM, supplemented with 10% (v/v) FCS, 1% (v/v) pen/strep and 1% (v/v) L-glutamine (2 mM final concentration, Gibco). All cell lines were maintained at 37°C and 5% CO2.

SARS-CoV-2 viral culture

VeroTMPRSS2 cells were seeded into 12 well plates one day prior to infection to be roughly 70% confluent. Growth media was removed from the VeroTMPRSS2 cells and 100µl of participant sample was added to each well. Plates were incubated at 37°C and 5% CO2 for 1 hour (h) with rocking every 10-15 min. After the inoculation period, 900µl Vero growth media was added to the cells and they were returned to the incubator for a 48h infection period. After 24h of that infection period, A549-ACE2 cells were seeded to 96 well black microplates with clear bottom (Corning CLS3603) at a density of 10,000 cells/well and cultured for the remaining 24 h of the infection of VeroTMPRSS2 cells. At the end of the 48h VeroTMPRSS2 infection, growth media was removed from the A549-ACE2 cells and 100µl of media from the infected VeroTMRPSS2 cells was transferred to the A549-ACE2 cells. These cells were then returned to the incubator for a 24h infection. Following infection, media were removed from wells and cells were fixed with neutral buffered formalin solution (10% Sigma HT501128) for at least 1h at 4°C prior to removal from the biosafety level 3 (BSL3) containment facility. All work with SARS-CoV-2 was performed under BSL3 containment at the University of Maryland, Baltimore following standard operating procedures.

Immunofluorescence staining and microscopy

For immunofluorescence staining, formalin solution was removed from the fixed cells as per lab standard procedures, then washed 1x with phosphate buffered saline (PBS) and any remaining formaldehyde was quenched with 50 mM $NH_4Cl$ in PBS, incubated for 15 minutes (min) at room temperature (as all subsequent steps). Following quenching, cells were washed 1x with PBS and permeabilized by 10 min incubation with 0.1% (v/v) Triton X-100 (Sigma) in 0.2% (w/v) bovine serum albumin (BSA, Sigma) solution in PBS (BSA/PBS). Samples were blocked by 10 min incubation with 0.2% BSA/PBS prior to addition of primary antibody. Anti-SARS-CoV-2 nucleocapsid (Sino Biological 40143-R004) was diluted 1:3000 in 0.2% BSA/PBS and incubated with samples for 1 h. Cells were washed 3x 5 min with 0.2% BSA/PBS to remove primary antibody then incubated with the secondary antibody (anti-rabbit IgG Alexa Fluor 488; Invitrogen A11008) used at a final concentration of 4 µg/ml in 0.2% BSA/PBS. Cells were washed 3x 5 min with PBS and then incubated with Hoechst 33342 (Invitrogen H3570) at a final concentration of 5 µg/ml (diluted in PBS) for 10 min for visualization of nuclei. Hoechst solution was removed from cells which were washed 1x with PBS then left in PBS to prevent drying of sample. Imaging was performed with a Celigo Imaging Cytometer (Nexcelom) for detection of all cells by Hoechst stain and infected cells by staining for the nucleocapsid protein labelling.

Antibody response assessment

Plasma samples were analyzed for antibodies to SARS-CoV-2 using a modified protocol described by Stadlbauer et al., 2020[3]. Samples were screened for IgG, IgM, and IgA antibodies to full length SARS-CoV-2 spike protein (produced by the Bioexpression and Fermentation Facility, University of Georgia)[4,5]. IgG and IgM antibodies to SARS-CoV-2 were confirmed and titered using the SARS-CoV-2 receptor binding domain (RBD) as the target[6].

ELISA plates were pre-coated overnight with full-length spike or RBD. Plates were washed, blocked, and washed again before screening at a 1:100 dilution, or confirmation and titering using four-fold serial dilutions (starting at 1:100 for IgG or 1:40 for IgM) of plasma samples. Diluted samples were added to the ELISA plates and incubated for one hour. The wells were then washed, incubated for one hour with horseradish peroxidase conjugated goat-anti-human IgA, IgM, and IgG, IgG or IgM detection antibody (Invitrogen), washed, incubated with 3,3′,5,5′-tetramethylbenzidine substrate (Seracare) for 10 minutes in the dark, and quenched with 1N sulfuric acid (Thermo Fisher). Plates were read at 450 nm. Positive and negative controls were run on each plate. Samples were considered positive if samples were above the established cut-offs on both tests for initial screening dilution and the first titer dilution.

SARS-CoV-2 sequencing and genome assembly

SARS-COV-2 RNA samples from saliva or nasal mid-turbinate swabs were subjected to whole-genome amplification by one-step RT-PCR with 39 primer pairs covering the 29,903 bp SARS-COV-2 reference genome (NC_045512.2) using the Access Array System, IFC controller AX and Fluidigm FC1 cycler (Fludigm Corporation, CA, USA)[7]. One well of integrated fluidic circuit (IFC) chip reaction consisted of 1.45 µl of RNA sample, 3 µl of 2x reaction mix, 0.2 µl of DMSO, 0,05 µl of RNase OUT, 0.05 µl of Superscript RT III high Fidelity Plantinum Taq polymerase, and 0.25 µl of 20x Access Array loading reagent. The primed IFC chip was loaded into the Fluidigm FC1 cycler and RT-PCR amplification were performed with the following cycle conditions: 30 min at 50 °C, 2 min at 94 °C, 40 cycles of 30 s at 94 °C, 30 s at 53 °C, 2 min at 68 °C, and 7 min at 68 °C. The RT-PCR products were purified using AMPure XP beads (Beckman coulter, CA) according to the manufactures protocol. The quality and quantity of the amplicons were determined by 4200 TapeStation system and DNA 5000 kit (Agilent Technologies, Waldbronn, Germany). The NGS libraries were prepared using Nextera DNA Flex Library Prep kit and Illumina DNA Unique Dual (UD) indexes (Illumina, CA, USA). Libraries were purified, and the fragment sizes and quality were analyzed by Agilent 4200 TapeStation system. Each library was pooled with equal molar ratio. Pooled library was normalized to 4 nM concentration and denatured with 5 µl of 0.2 N sodium hydroxide. The 14 pM library was spiked with 1% PhiX control (PhiX Control v3) and sequenced on an Illumina MiSeq platform using a MiSeq reagent kit v3 ( 600 cycles). Fastqs for each of the samples were ran on ngs_mapper genome assembly pipeline[8] for data cleaning through Trimmomatic, and for reference-based genome assembly to NC_045512.2 SARS-CoV-2 reference genome through BWA-MEM. Variant calling was performed with a minimum Phred of 25 and ambiguous bases called when present at a frequency of 20% or higher, followed by manual curation of the assembled genomes for removal of primer induced error. SARS-COV-2 lineages and mutations were determined using PANGOLIN and NextStrain tools[9,10]. All single nucleotide variant calls specific to variants of concern (VOC) were validated by mapping the reads for each sample back to the assembled consensus genome using BWA-MEM[11] and inspecting coverage plots with IGV[12] and allele frequencies via SAMtools mpileup[13] and LoFreq[14].

Additional statistical analysis methods

The probability of culture positive and SARS-CoV-2 RNA copy numbers detected by qRT-PCR in mid-turbinate swab and saliva samples was estimated based on a logistic regression model with the status of culture positivity as the outcome and the log-10 transformed RNA copy numbers as the independent variable, using the glm function from the R package “stats.” Standard errors from the logistic model were used to estimate the confidence interval around the regression line (Figure S2).

**Supplemental Tables**

**Table S1.** Variant Classification

| **Variant** | **Antibody negative at enrollment N (%)** | **Antibody positive at enrollment N (%)** | **Antibody positive and negative N (%)** |
|---|---|---|---|
| **Wild type or other** | 36 (73) | 4 (50) | 40 (70) |
| **Alpha** | 4 (8) | 0 (0) | 4 (7) |
| **Gamma** | 2 (4) | 0 (0) | 2 (4) |
| **Iota** | 1 (2) | 1 (12) | 2 (4) |
| **Undetermined/unknown** | 6 (12) | 3 (38) | 9 (16) |
| **Total** | 49 (100) | 8 (100) | 57 (100) |

**Table S2A.** Viral RNA copies and culture of samples from cases seronegative for SARS-CoV-2 antibodies at the first shedding assessment.

| Sample Type | Cases[a] | Cases with ≥1 positive sample[b] N (%) | | Samples | Positive Samples[c] N (%) | | Culture n/N (%)[d] | GM (95% CI) [e] | GSD | Maximum RNA copies[f] |
|---|---|---|---|---|---|---|---|---|---|---|
| | | ≥LOD | ≥LOQ | | ≥LOD | ≥LOQ | | | | |
| **Mid-turbinate Swab** | 49 | 49 (100) | 49 (100) | 80 | 80 (100) | 79 (99) | 50/73 (68) | $5.7 \times 10^6$ ($2.1 \times 10^6$, $1.5 \times 10^7$) | 1.2 | $5.1 \times 10^9$ |
| **Saliva** | 49 | 48 (98) | 48 (98) | 80 | 79 (99) | 76 (95) | 20/62 (32) | $2.7 \times 10^5$ ($1.1 \times 10^5$, $6.9 \times 10^5$) | 1.3 | $5.2 \times 10^8$ |
| **Fomite** | 49 | 32 (65) | 18 (37) | 80 | 42 (52) | 21 (26) | 0/73 (0) | 64 (26, 160) | 1.4 | $1.2 \times 10^6$ |
| **Coarse Aerosol without mask** | 49 | 15 (31) | 7 (14) | 78 | 20 (26) | 8 (10) | 0/38 (0) | 6.2 (2.4, 16) | 2 | $5.1 \times 10^4$ |
| **Coarse Aerosol with mask** | 46 | 10 (22) | 2 (4) | 71 | 13 (18) | 2 (3) | 0/16 (0) | 15 (9.7, 22) | 1.8 | $6.4 \times 10^3$ |
| **Fine Aerosol without mask** | 49 | 22 (45) | 11 (22) | 78 | 28 (36) | 13 (17) | 0/75 (0) | 19 (8.6, 41) | 1.8 | $5.4 \times 10^4$ |
| **Fine Aerosol with mask** | 46 | 14 (30) | 3 (7) | 71 | 17 (24) | 3 (4) | 2/66 (3) | 7.6 (3.4, 17) | 1.9 | $2.0 \times 10^4$ |

[a] Participants with MTS or saliva samples positive for SARS-CoV-2 viral RNA by qRT-PCR and seronegative for SARS-CoV-2 spike protein antibody at their first breath sampling visit, and who provided at least one 30-minute exhaled breath sample.

[b] Number of participants with at least one sample ≥LOD or ≥LOQ as described below.

[c] Samples positive and ≥LOD had at least one replicate with confirmed amplification after inspection and quality control (LOD ~75 RNA copies with 95% probability of detection) and ≥LOQ if the mean of replicate assays was ≥ 250 RNA copies.

[d] n/N = Number of positive cultures/Number of samples cultured (percent positive).

[e] GM = geometric mean; GSD = geometric standard deviation. The GM and GSD were computed, accounting for samples below the LOD, using a linear mixed-effects model for censored responses (R Project package "lmec") using data for all samples of each sample type with nested random effects of samples within study participant.

[f] The largest quantity of RNA copies detected based on the mean from replicates qRT-PCR aliquots.

**Table S2B.** Viral RNA copies and culture of samples from alpha variant cases, seronegative for SARS-CoV-2 antibodies at the first assessment.

| Sample Type | Cases[a] | Cases with ≥1 positive sample[b] N (%) | | Samples | Positive Samples[c] N (%) | | Culture n/N (%) [d] | GM (95% CI)[e] | GSD | Maximum RNA copies[f] |
|---|---|---|---|---|---|---|---|---|---|---|
| | | ≥LOD | ≥LOQ | | ≥LOD | ≥LOQ | | | | |
| **Mid-turbinate Swab** | 4 | 4 (100) | 4 (100) | 6 | 6 (100) | 6 (100) | 6/6 (100) | $3.8 \times 10^8$ ($3.3 \times 10^8$, $4.4 \times 10^8$) | 1.2 | $2.9 \times 10^9$ |
| **Saliva** | 4 | 4 (100) | 4 (100) | 6 | 6 (100) | 6 (100) | 3/5 (60) | $1.9 \times 10^7$ ($2.7 \times 10^6$, $1.3 \times 10^8$) | 1.2 | $5.2 \times 10^8$ |
| **Fomite** | 4 | 4 (100) | 4 (100) | 6 | 6 (100) | 4 (67) | 0/6 (0) | 560 (530, 590) | 1.1 | $1.7 \times 10^4$ |
| **Coarse Aerosol without mask** | 4 | 4 (100) | 2 (50) | 6 | 6 (100) | 3 (50) | 0/6 (0) | 140 (28, 730) | 2.2 | $5.1 \times 10^4$ |
| **Coarse Aerosol with mask** | 4 | 3 (75) | 2 (50) | 6 | 4 (67) | 2 (33) | 0/5 (0) | 29 (23, 36) | 1.2 | $6.4 \times 10^3$ |
| **Fine Aerosol without mask** | 4 | 4 (100) | 3 (75) | 6 | 6 (100) | 4 (67) | 0/6 (0) | 580 (450, 740) | 1.4 | $5.4 \times 10^4$ |
| **Fine Aerosol with mask** | 4 | 4 (100) | 3 (75) | 6 | 5 (83) | 3 (50) | 1/5 (20) | 290 (180, 470) | 1.6 | $2.0 \times 10^4$ |

[a] Participants (alpha variant cases) with MTS or saliva samples positive for SARS-CoV-2 viral RNA by qRT-PCR and seronegative for SARS-CoV-2 spike protein antibody at their first breath sampling visit, and who provided at least one 30-minute exhaled breath sample.

[b] Number of participants with at least one sample ≥LOD or ≥LOQ as described below.

[c] Samples positive and ≥LOD had at least one replicate with confirmed amplification after inspection and quality control (LOD ~75 RNA copies with 95% probability of detection) and ≥LOQ if the mean of replicate assays was ≥ 250 RNA copies.

[d] n/N = Number of positive cultures/Number of samples cultured (percent positive).

[e] GM = geometric mean; GSD = geometric standard deviation. The GM and GSD were computed, accounting for samples below the LOD, using a linear mixed-effects model for censored responses (R Project package "lmec") using data for all samples of each sample type with nested random effects of samples within study participant.

[f] The largest quantity of RNA copies detected based on the mean from replicates qRT-PCR aliquots.

**Table S2C.** Viral RNA copies and culture of samples from wild type and other (non-alpha) variant cases, seronegative for SARS-CoV-2 antibodies at the first assessment.

| Sample Type | Cases[a] | Cases with ≥1 positive sample[b] N (%) | | Samples | Positive Samples[c] N (%) | | Culture n/N (%) [d] | GM (95% CI)[e] | GSD | Maximum RNA copies[f] |
|---|---|---|---|---|---|---|---|---|---|---|
| | | ≥LOD | ≥LOQ | | ≥LOD | ≥LOQ | | | | |
| **Mid-turbinate Swab** | 45 | 45 (100) | 45 (100) | 74 | 74 (100) | 73 (99) | 44/67 (66) | $3.8 \times 10^6$ ($1.4 \times 10^6$, $1.0 \times 10^7$) | 1.2 | $5.1 \times 10^9$ |
| **Saliva** | 45 | 44 (98) | 44 (98) | 74 | 73 (99) | 70 (95) | 17/57 (30) | $2.1 \times 10^5$ ($8.1 \times 10^4$, $5.4 \times 10^5$) | 1.3 | $3.9 \times 10^8$ |
| **Fomite** | 45 | 28 (62) | 14 (31) | 74 | 36 (49) | 17 (23) | 0/67 (0) | 46 (17, 120) | 1.5 | $1.2 \times 10^6$ |
| **Coarse Aerosol without mask** | 45 | 11 (24) | 5 (11) | 72 | 14 (19) | 5 (7) | 0/32 (0) | 7 (3.2, 15) | 1.7 | $2.6 \times 10^4$ |
| **Coarse Aerosol with mask** | 42 | 7 (17) | - | 65 | 9 (14) | 0 (0) | 0/11 (0) | 15 (10, 21) | 1.9 | 200 |
| **Fine Aerosol without mask** | 45 | 18 (40) | 8 (18) | 72 | 22 (31) | 9 (12) | 0/69 (0) | 18 (9.1, 34) | 1.9 | $2.8 \times 10^3$ |
| **Fine Aerosol with mask** | 42 | 10 (24) | - | 65 | 12 (18) | 0 (0) | 1/61 (2) | 24 (18, 31) | 1.8 | 260 |

[a] Participants (wild type and non-alpha variant cases) with MTS or saliva samples positive for SARS-CoV-2 viral RNA by qRT-PCR and seronegative for SARS-CoV-2 spike protein antibody at their first breath sampling visit, and who provided at least one 30-minute exhaled breath sample.

[b] Number of participants with at least one sample ≥LOD or ≥LOQ as described below.

[c] Samples positive and ≥LOD had at least one replicate with confirmed amplification after inspection and quality control (LOD ~75 RNA copies with 95% probability of detection) and ≥LOQ if the mean of replicate assays was ≥ 250 RNA copies.

[d] n/N = Number of positive cultures/Number of samples cultured (percent positive).

[e] GM = geometric mean; GSD = geometric standard deviation. The GM and GSD were computed, accounting for samples below the LOD, using a linear mixed-effects model for censored responses (R Project package "lmec") using data for all samples of each sample type with nested random effects of samples within study participant.

[f] The largest quantity of RNA copies detected based on the mean from replicates qRT-PCR aliquots

**Table S3.** The effect of alpha variant on viral load of Mid-turbinate swab and Saliva

| | **Mid-turbinate swab**<br>**N=49**<br>**n=80[a]**<br>**median = 1.0 x $10^7$**<br>**IQR (2.7 x $10^5$, 1.1 x $10^8$)** | **Saliva**<br>**N=49**<br>**n=80**<br>**median = 3.6 x $10^5$**<br>**IQR (2.9 x $10^4$, 1.9 x $10^6$)** |
|---|---|---|
| | **Unadjusted Estimates**<br>**(95% CI)** | **Unadjusted Estimates**<br>**(95% CI)** |
| **Alpha variant** | 170 (5.4, 5300) | 35 (1.2, 980) |
| **Age** | 1.3 (0.44, 3.6) | 2.2 (0.82, 5.6) |
| **Day post-symptom onset** | 0.82 (0.57, 1.2) | 0.84 (0.61, 1.1) |
| **Log mid-turbinate swab** | - | 9.7 (3.6, 26) |
| **Log saliva** | 9.2 (3.7, 23) | - |
| **Gastrointestinal symptoms** | 3.6 (1.5, 8.5) | 1.7 (0.79, 3.5) |
| **Lower respiratory symptoms** | 1.5 (0.55, 3.9) | 1.6 (0.65, 3.7) |
| **Systemic symptoms** | 4.6 (1.8, 12) | 3.1 (1.4, 6.9) |
| **Upper respiratory symptoms** | 5 (2.1, 12) | 2.6 (1.3, 5.4) |

Effect estimates and their 95% confidence intervals are shown as the RNA copy number ratio of alpha to non-alpha variant, or as the fold-increase in RNA copy number for a 10-year increase in age, 1-day increase in day post-symptom onset, and an inner-quartile range change in symptom scores, MTS, and saliva RNA copy number. All analyses were controlled for random effects of subject, and sample nested within subjects, using a linear mixed-effects model for censored responses (R Project package "lmec") by the limit of detection.

[a] N = Number of participants, n = Number of samples

**Table S4.** Frequency of different mask types at the sample level used in the paired analyses

| | With mask | | | Without mask |
|---|---|---|---|---|
| | **Cloth mask or double mask** | **Surgical mask** | **KN95** | |
| **Fine and Coarse Aerosols** | **22 (32%)** | **46 (67%)** | **1 (1%)** | **69** |

**Table S5.** Regression Coefficients for Effect of Masks and Mask Type

| | **Coarse Aerosol** | **Fine Aerosol** |
|---|---|---|
| **Intercept** | 2.3 (1.5, 3) | 2.8 (2, 3.6) |
| **Any mask** | -0.84 (-1.7, 0.019) | -1.5 (-2.4, -0.69) |
| **Surgical mask** | 0.09 (-0.99, 1.2) | 0.72 (-0.28, 1.7) |
| **Cough** | 0.13 (0.0018, 0.26) | 0.11 (-0.019, 0.24) |

Regression to identify the relative performance of cloth masks and surgical masks, dropping the one observation made on a person wearing a KN95.

**Table S6.** Viral RNA copy numbers in participants who were already seropositive at enrollment.

| Sample Type | Cases[a] | Cases with ≥1 positive sample[b] N (%) | | Samples | Positive Samples[c] N (%) | | Culture n/N (%)[d] | GM (95% CI)[e] | GSD | Maximum RNA copies[f] |
|---|---|---|---|---|---|---|---|---|---|---|
| | | ≥LOD | ≥LOQ | | ≥LOD | ≥LOQ | | | | |
| Mid-turbinate Swab | 8 | 7 (88) | 7 (88) | 15 | 13 (87) | 13 (87) | 6/11 (55) | $1.6 \times 10^5$ ($9.5 \times 10^3$, $2.9 \times 10^6$) | 1.3 | $8.4 \times 10^8$ |
| Saliva | 8 | 8 (100) | 7 (88) | 15 | 15 (100) | 13 (87) | 1/11 (9) | $2.2 \times 10^4$ ($5.4 \times 10^3$, $9.0 \times 10^4$) | 1.3 | $2.3 \times 10^7$ |
| Phone Swab | 8 | 3 (38) | 2 (25) | 15 | 4 (27) | 2 (13) | 0/11 (0) | 17 (4.4, 66) | 1.5 | 600 |
| Coarse Aerosol without mask | 8 | 0 (0) | 0 (0) | 15 | 0 (0) | 0 (0) | 0/2 (0) | - | - | - |
| Coarse Aerosol with mask | 8 | 0 (0) | 0 (0) | 14 | 0 (0) | 0 (0) | 0/2 (0) | - | - | - |
| Fine Aerosol without mask | 8 | 0 (0) | 0 (0) | 15 | 0 (0) | 0 (0) | 0/13 (0) | - | - | - |
| Fine Aerosol with mask | 8 | 0 (0) | 0 (0) | 14 | 0 (0) | 0 (0) | 0/13 (0) | - | - | - |

[a] Participants with a mid-turbinate or saliva samples positive for SARS-CoV-2 viral RNA by qRT-PCR and seropositive for SARS-CoV-2 spike protein antibody at enrollment and who provided at least one 30-minute sample of exhaled breath.

[b] Number of participants with at least one sample ≥LOD or ≥LOQ as described below.

[c] Samples positive and ≥LOD had at least one replicate with confirmed amplification after inspection and quality control (LOD ~75 RNA copies with 95% probability of detection) and ≥LOQ if the mean of replicate assays was ≥ 250 RNA copies.

[d] n/N = Number of positive cultures/Number of samples cultured (percent positive).

[e] GM = geometric mean; GSD = geometric standard deviation. The GM and GSD were computed controlled for random effects of subject and sample nested within subjects using a linear mixed-effects model for censored responses (R Project package "lmec") by the limit of detection.

[f] The largest quantity of RNA copies detected based on the mean from replicate qRT-PCR aliquots.

**Table S7.** Probability of positive cultures at GM viral RNA copy numbers observed.

| Level | Sample Type | Variant | RNA copies | Probability based on mid-turbinate model | Probability based on saliva model |
|---|---|---|---|---|---|
| $TCID_{50}$ | Mid-turbinate swab | All | $7.8 \times 10^5$ | 0.5 | |
| | Saliva | All | $5.2 \times 10^6$ | | 0.5 |
| $TCID_{63}$ | Mid-turbinate swab | All | $2.0 \times 10^6$ | 0.63 | |
| | Saliva | All | $2.2 \times 10^7$ | | 0.63 |
| Geometric mean level | Mid-turbinate swab | Total | $5.7 \times 10^6$ | 0.76 | 0.51 |
| | | Alpha | $3.8 \times 10^8$ | 0.97 | 0.84 |
| | | Other | $3.8 \times 10^6$ | 0.71 | 0.47 |
| | Saliva | Total | $2.7 \times 10^5$ | 0.35 | 0.24 |
| | | Alpha | $1.9 \times 10^7$ | 0.86 | 0.62 |
| | | Other | $2.1 \times 10^5$ | 0.32 | 0.23 |
| | Fomite | Total | 64 | 0.0046 | 0.013 |
| | | Alpha | 560 | 0.016 | 0.03 |
| | | Other | 46 | 0.0038 | 0.012 |
| | Coarse (> 5 µm) Aerosol | Total | 6.2 | 0.0012 | 0.0055 |
| | | Alpha | 140 | 0.0072 | 0.018 |
| | | Other | 7 | 0.0013 | 0.0057 |
| | Fine (≤ 5 µm) Aerosol | Total | 19 | 0.0023 | 0.0084 |
| | | Alpha | 580 | 0.016 | 0.03 |

| | | | | | |
|---|---|---|---|---|---|
| | | Other | 18 | 0.0022 | 0.0082 |
| Maximum RNA copies | Mid-turbinate swab | Alpha | $2.9 \times 10^9$ | 0.99 | 0.92 |
| | | Other | $5.1 \times 10^9$ | 0.99 | 0.93 |
| | Saliva | Alpha | $5.2 \times 10^8$ | 0.98 | 0.85 |
| | | Other | $3.9 \times 10^8$ | 0.97 | 0.84 |
| | Fomite | Alpha | $1.7 \times 10^4$ | 0.1 | 0.1 |
| | | Other | $1.2 \times 10^6$ | 0.56 | 0.36 |
| | Coarse (> 5 µm) Aerosol | Alpha | $5.1 \times 10^4$ | 0.17 | 0.15 |
| | | Other | $2.6 \times 10^4$ | 0.13 | 0.12 |
| | Fine (≤ 5 µm) Aerosol | Alpha | $5.4 \times 10^4$ | 0.18 | 0.15 |
| | | Other | $2.8 \times 10^3$ | 0.039 | 0.054 |

### Supplemental Figures

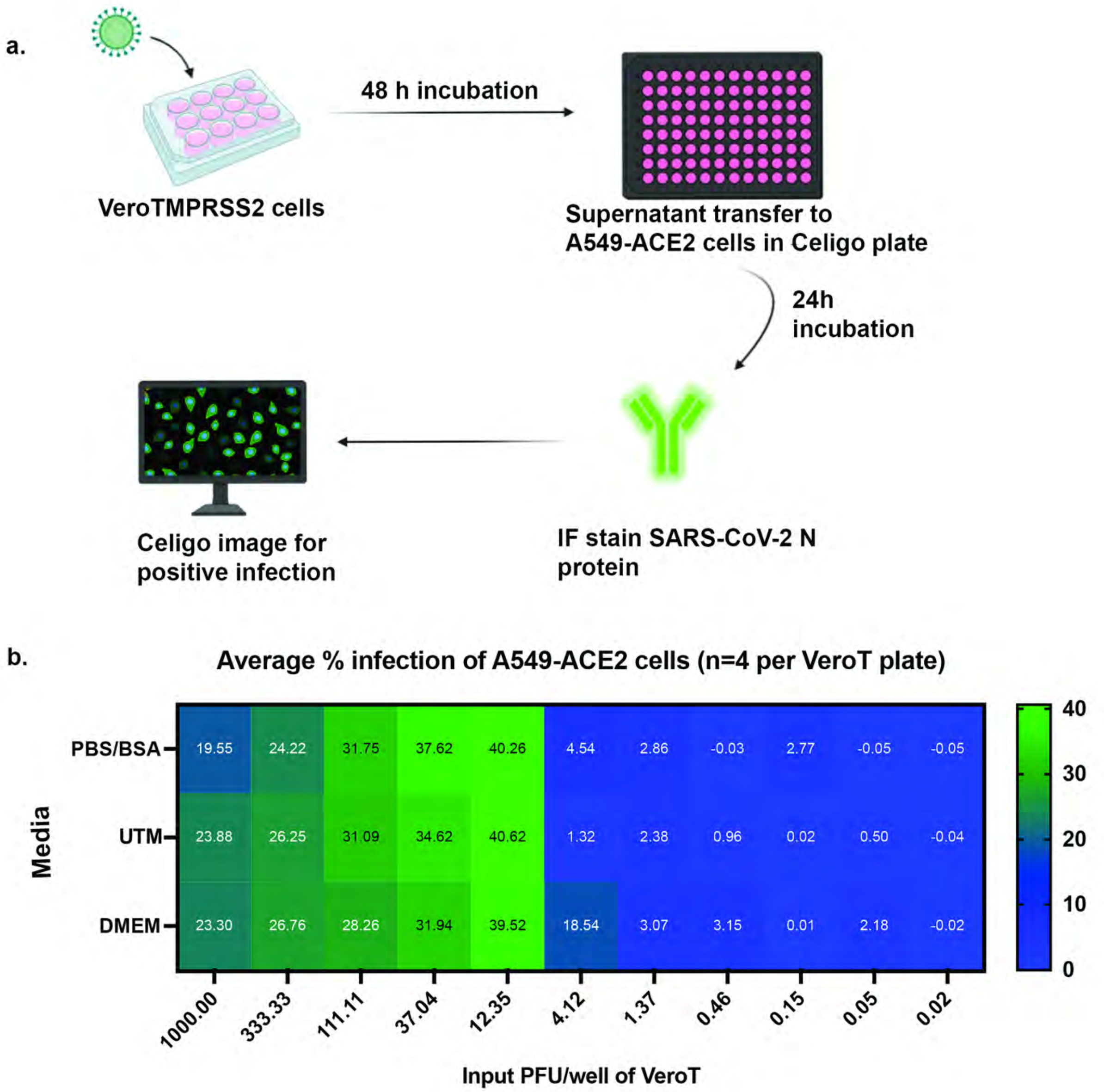

Figure S1. Culture methods and sensitivity

**a.** Diagram of the tissue culture amplification, focus assay, and visualization process. **b.** Limit of detection for SARS-CoV2 infection as tested using different sample collection media. Virus was diluted in PBS with 0.1% BSA, Universal Transport Media (UTM), or DMEM and added to VeroT cells. After amplification on the VeroT cells, supernatant was transferred to A549-ACE2 cells. Percent cells infected was measured by Fluorescent Focus Assay.

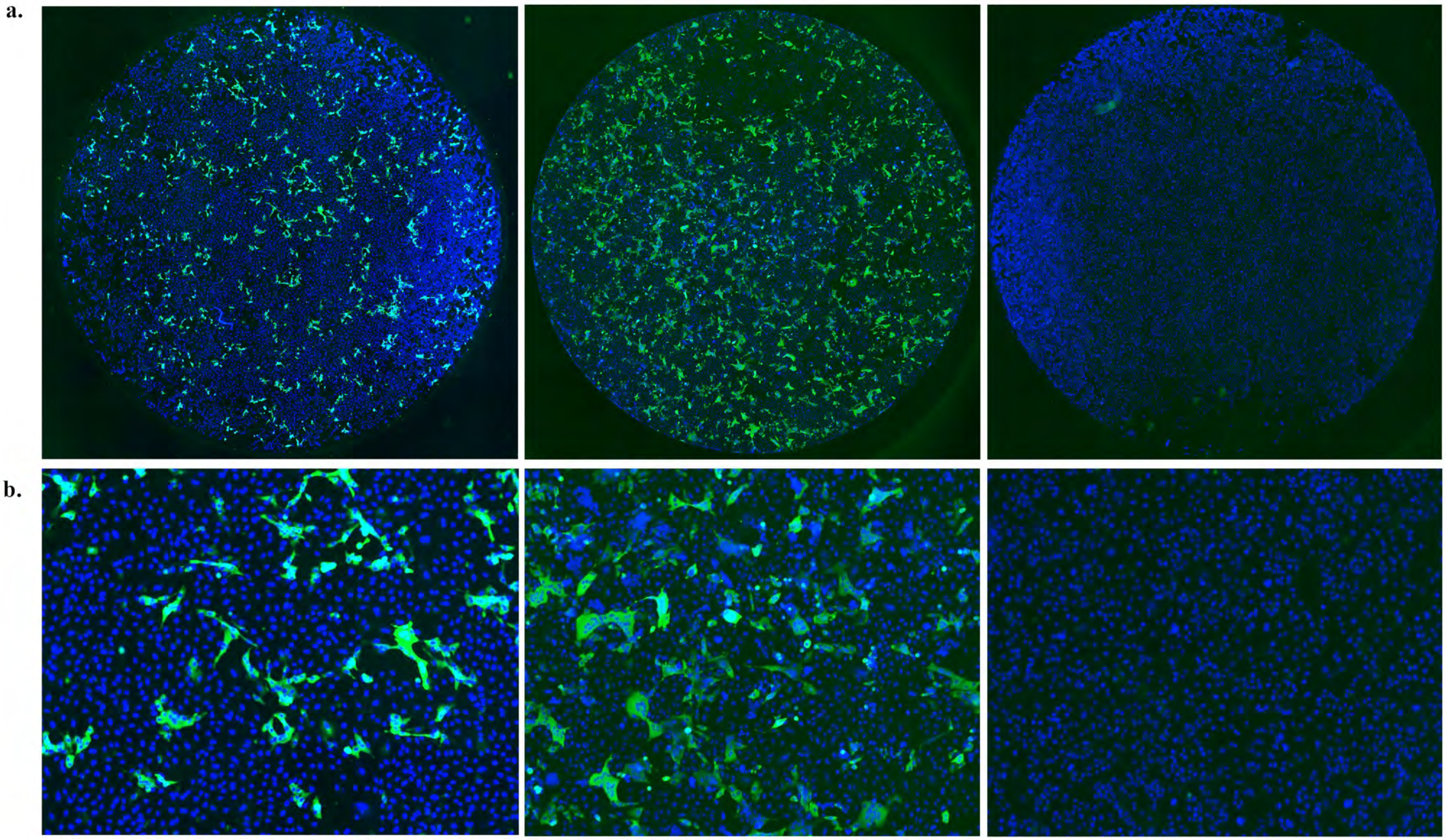

<u>Figure S2. Culture results from representative samples</u>
Example images from the focus assays as whole well images (**a**) and zoom in of the center of the well (**b**). Left, positive fine aerosol sample. Center, positive mid-turbinate swab sample. Right, negative fine aerosol sample.

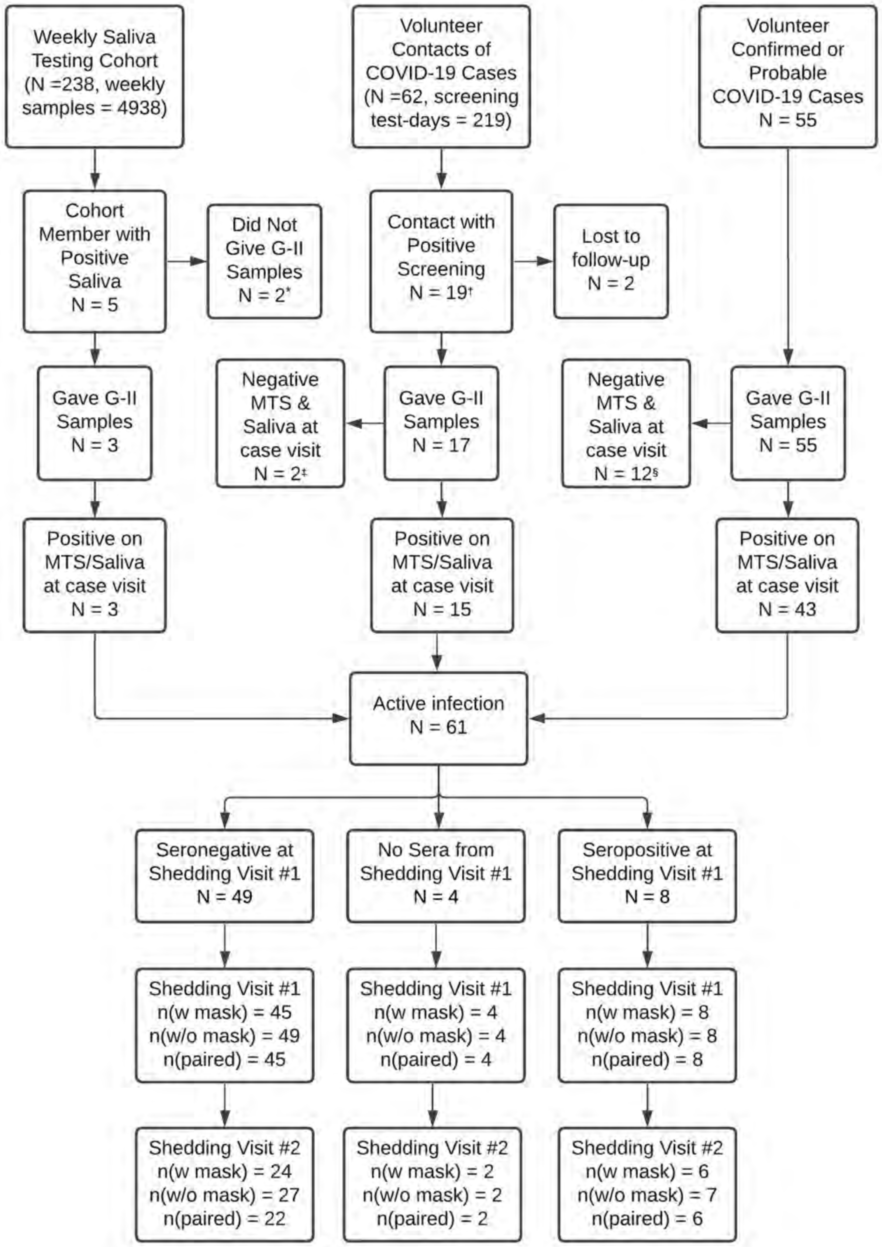

Figure S3. Study design

Study profile. Saliva screening was performed from May 8, 2020 through June 1, 2021. The first case to give a confirmed G-II was enrolled on June 6, 2020.

[a]One participant declined to enroll in a shedding visit and limited accessibility prevented the other from entering the G-II booth.

[b]Positive screening was determined by either PCR or symptoms.

[c]Two contacts had transient positive samples during repeated testing to identify early cases but were negative on further testing.

[d]Among volunteer cases with a clinical diagnosis of COVID-19, 12 were negative for viral RNA when seen for breath testing.

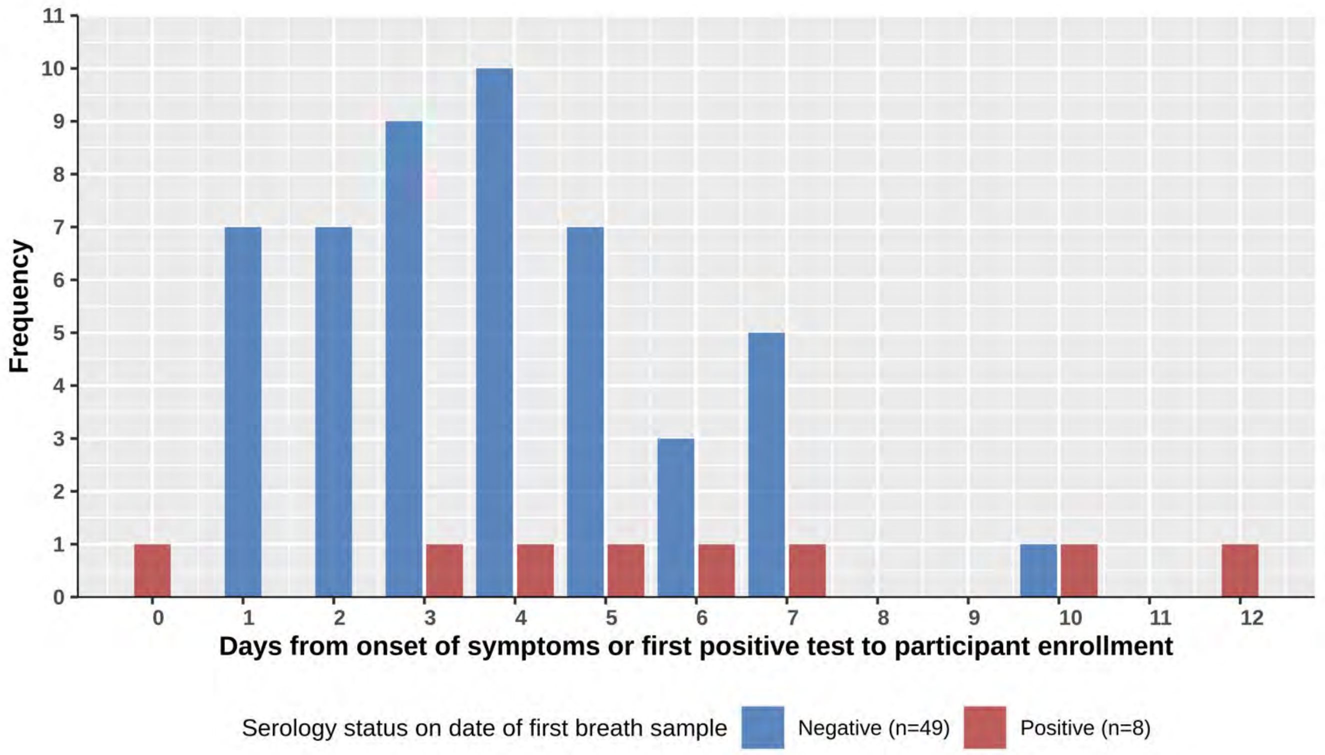

Figure S4. Days since onset
Comparison of the number of days from the date of onset of symptoms for symptomatic participants (or date of the first positive test in asymptomatic participants) to the day of their first breath sampling. 3 of the 49 subjects who were antibody-negative at enrollment had no symptoms, and they enrolled between two and four days after the day of their first positive results. Breath samples were collected on the day of enrollment in 46 participants, 1 day following enrollment in 3 participants, 2 days following enrollment in 3 participants, 3 days following enrollment in 2 participants, and 5 days following enrollment in 3 participants. The mean number of days (+/- SD) post symptom onset or since the first positive test was 5.9 +/- days in symptomatic participants who tested positive in their initial antibody screen; 3.8 +/- 2.1 days for symptomatic participants who tested negative; and 3.3 +/- 1.2 days in asymptomatic participants.

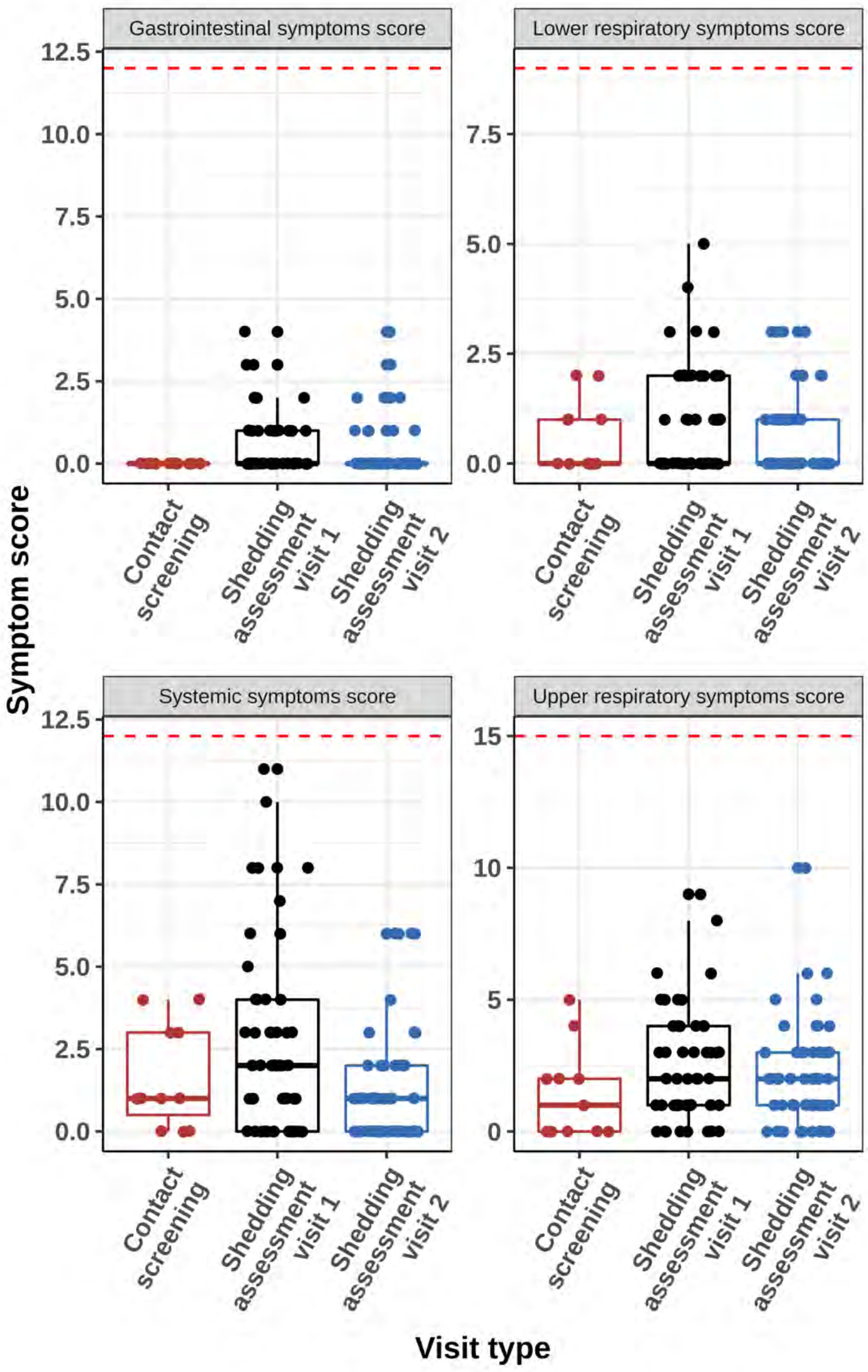

<u>Figure S5. Symptoms</u>
Symptoms reported during contact and shedding visits for case participants who tested seronegative at their first breath sample collection. The red dashed line is at the maximum score: systemic symptoms max score =12, gastrointestinal symptoms max score = 12, lower respiratory symptoms max score = 9, upper respiratory symptoms max score = 15.

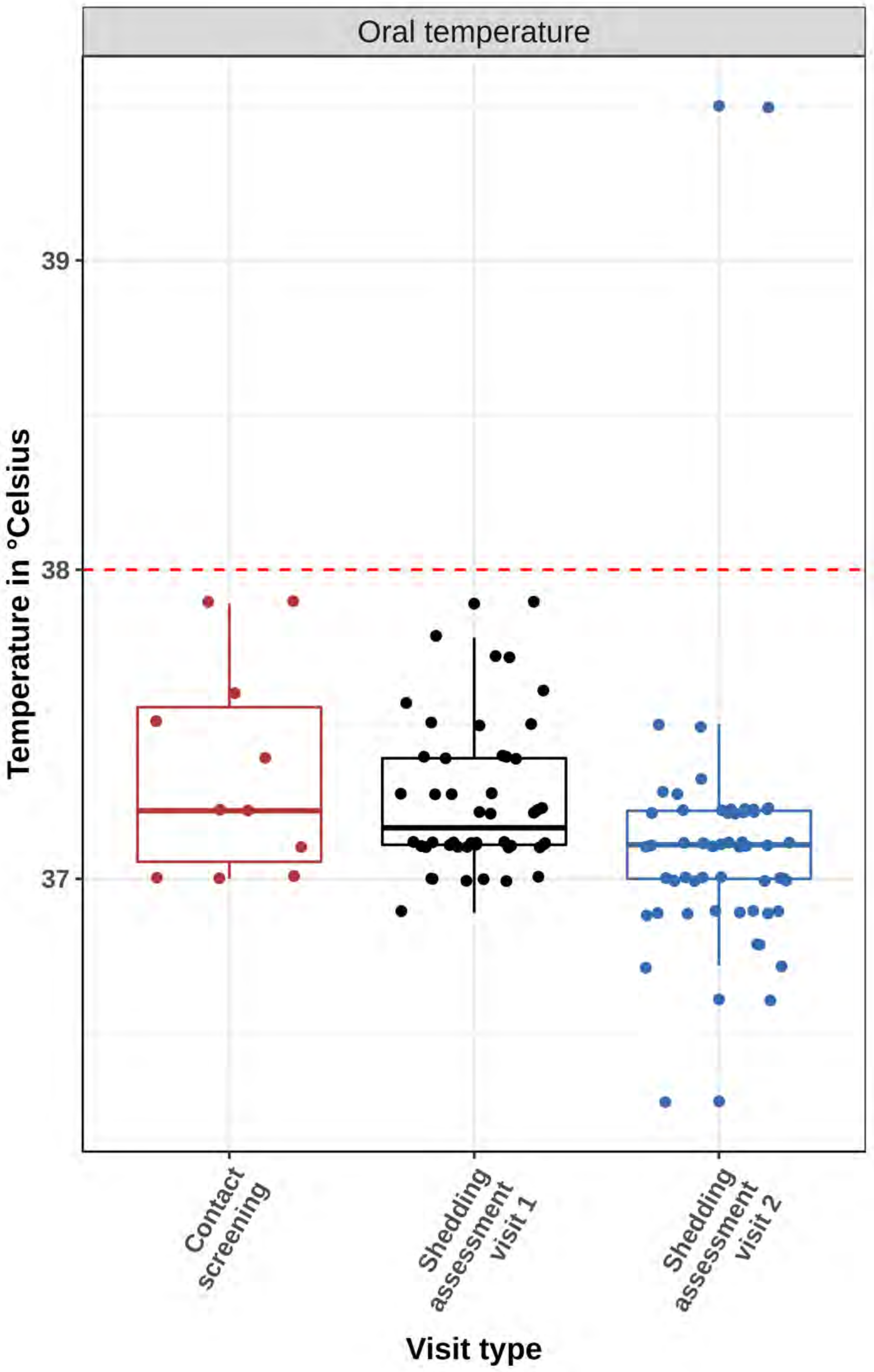

<u>Figure S6. Temperature</u>
Measured oral temperature at clinic visits of case participants who were seronegative at the time of first breath sample (N=49). The red dashed line is the fever threshold at 38 °C (100.4 °F).

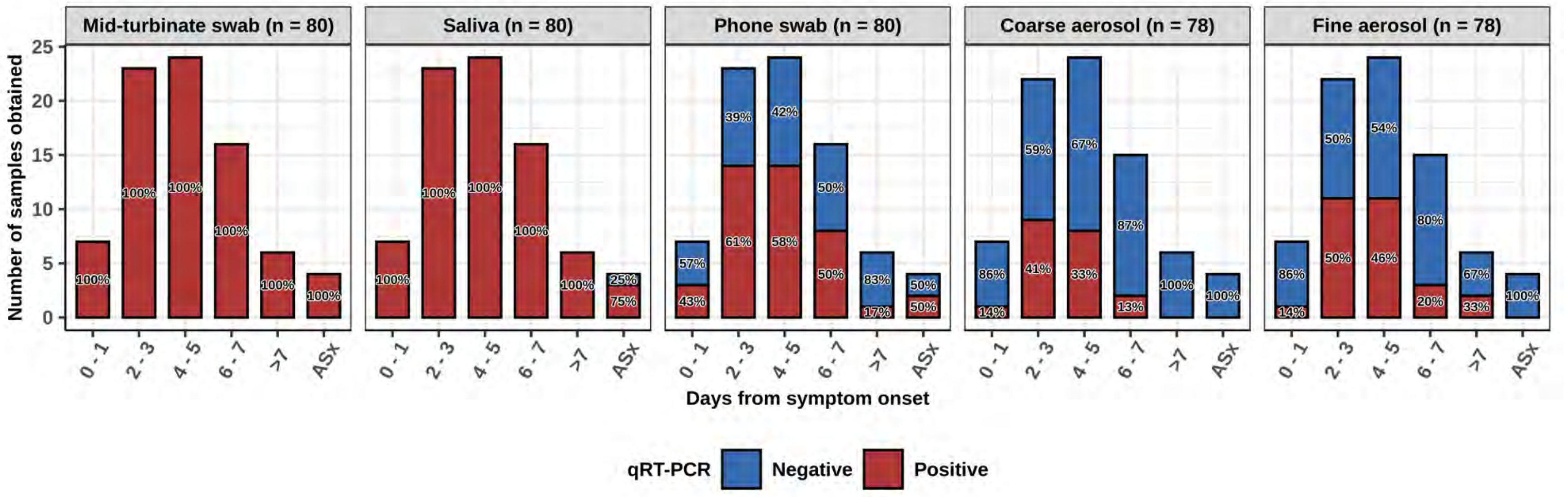

Figure S7. Positive and negative samples by day since onset.

Bar plots showing the number of samples tested and positive or negative status by RT-qPCR from study participants who were seronegative at first breath sample collection categorized by the approximate number of days post symptom onset (or first positive test for asymptomatic cases) when they were obtained. The percentages are percentage of each sample type with viral RNA above the limit of detection (red) or not detected (blue). Coarse and fine aerosols (without mask) were collected over a 30-minute sampling period. Asx = Asymptomatic.

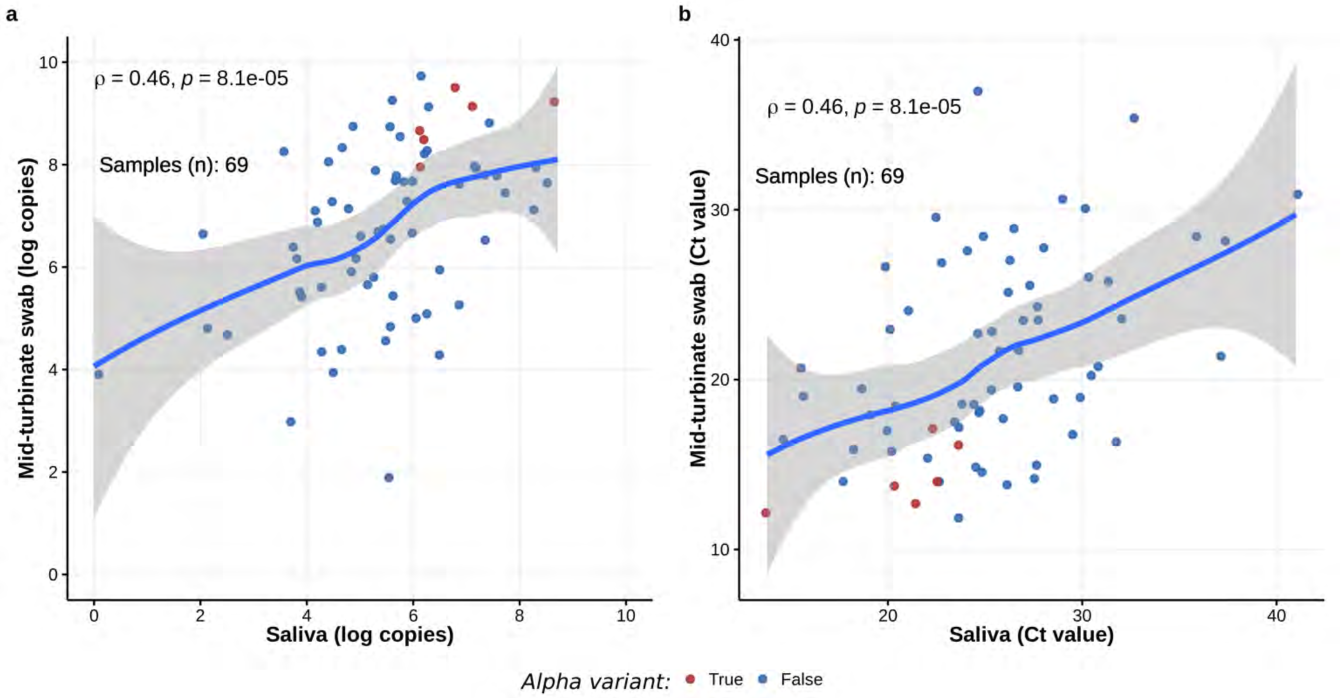

<u>Figure S8. Correlation between saliva and mid-turbinate swab RNA measurements</u>

**a.** Correlation between the SARS-CoV-2 RNA copy numbers detected by qRT-PCR in upper respiratory samples (saliva and mid-turbinate swab). **b.** Correlation between the cycle threshold (Ct) values for SARS-CoV-2 N-gene that was detected by qRT- PCR in the upper respiratory samples (saliva and mid-turbinate swab). ρ = *Spearman correlation coefficient.*

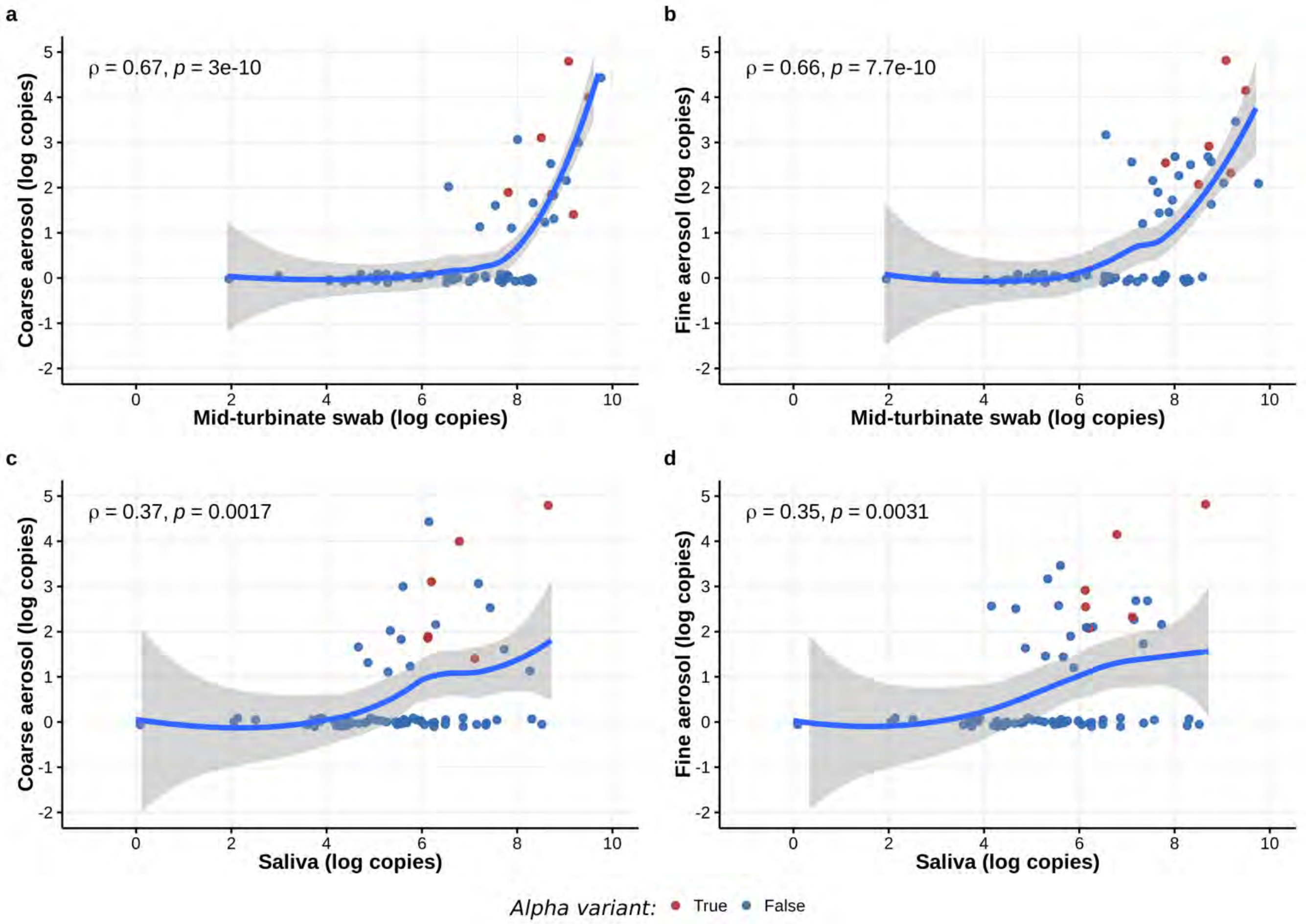

Figure S9. Correlation between the SARS-CoV-2 RNA copy numbers detected by qRT-PCR in upper respiratory samples (saliva and mid-turbinate swabs) and exhaled breath samples

Data shown are for samples collected without mask wearing. $\rho$ = *Spearman correlation coefficient.*

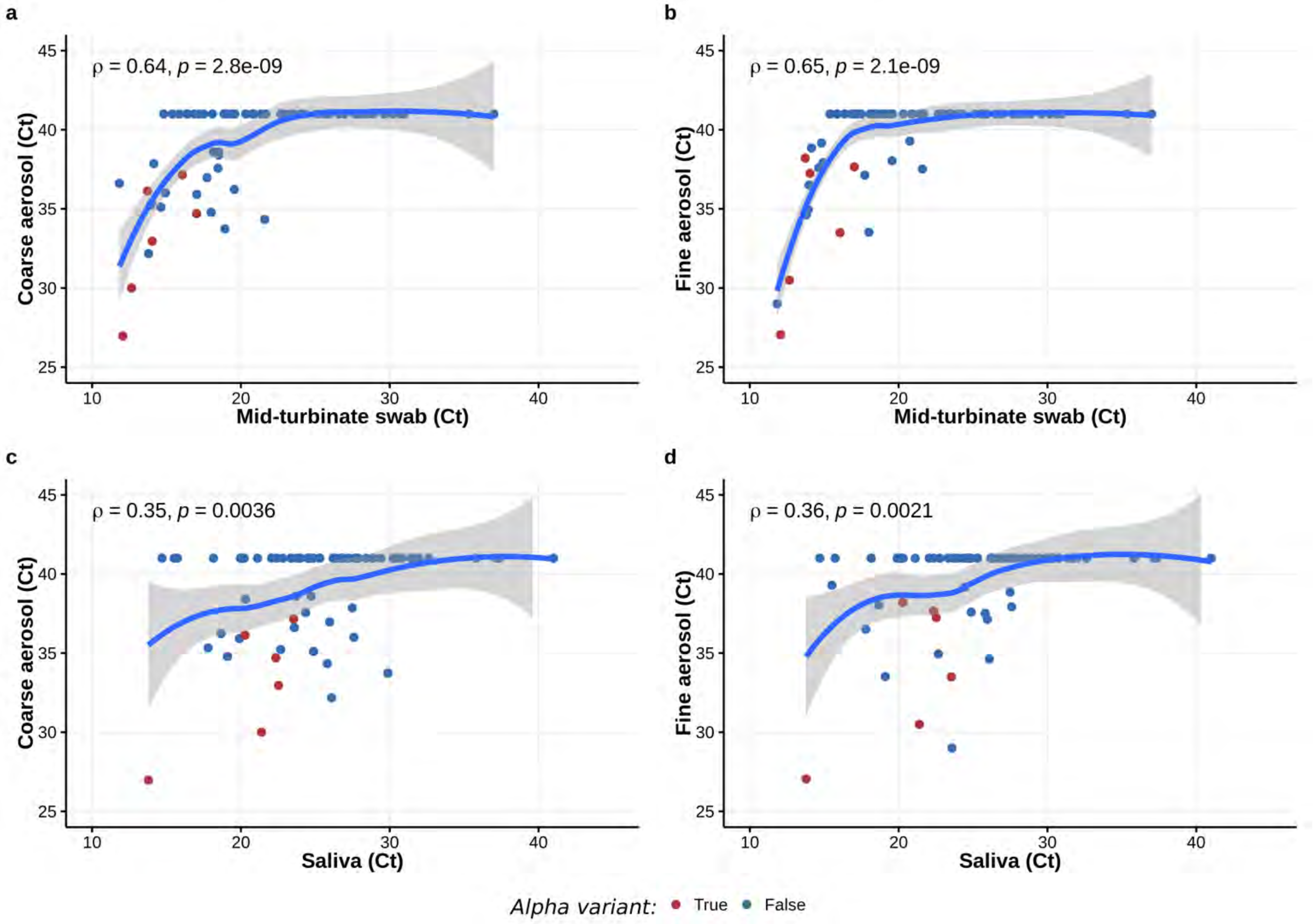

<u>Figure S10. Correlation between the cycle threshold (Ct) values for SARS-CoV-2 N-gene in the upper respiratory samples (saliva and mid-turbinate swabs) and exhaled breath samples.</u>

Data shown are for samples collected without mask wearing. $\rho$ = *Spearman correlation coefficient.*

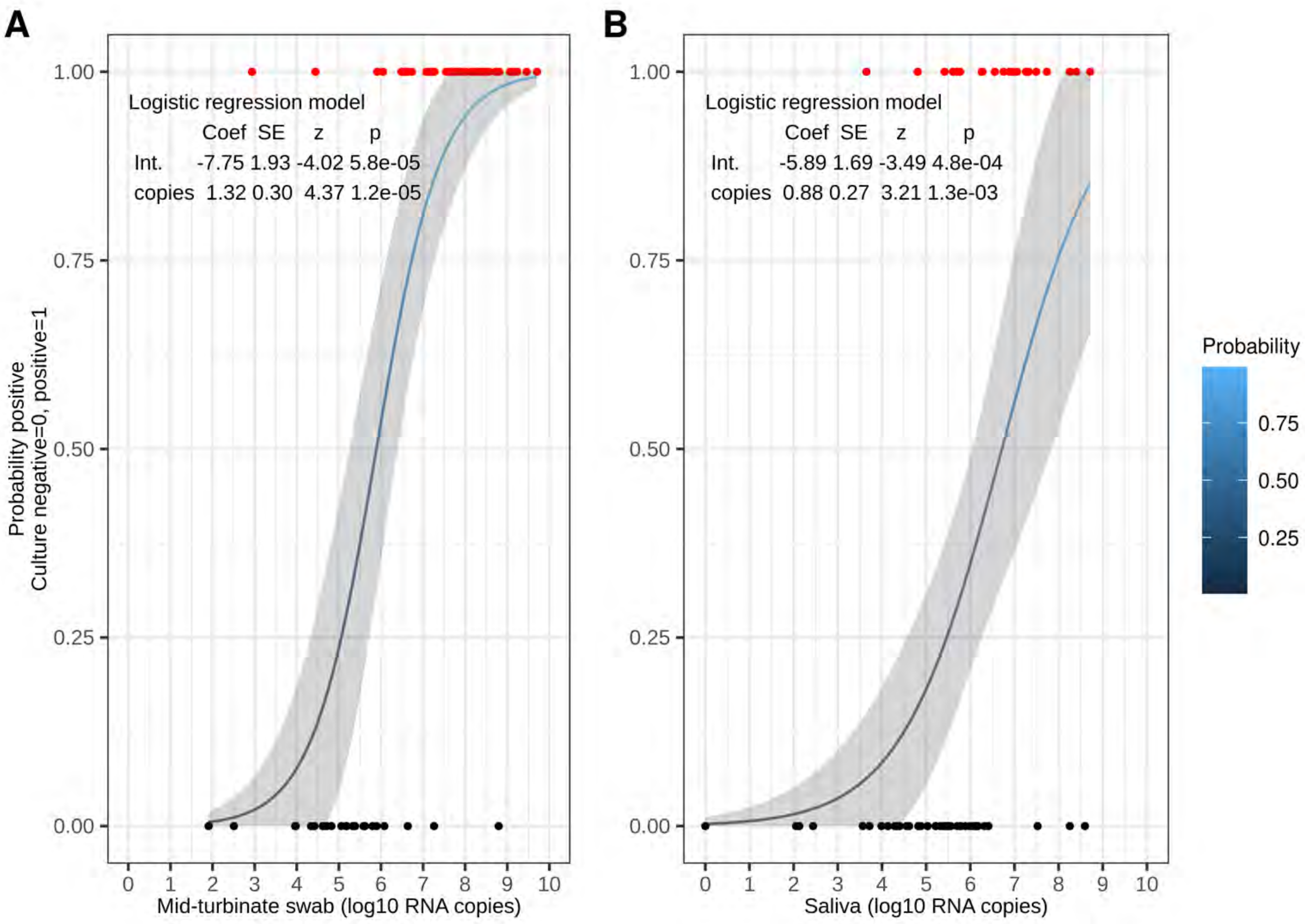

Figure S11. Probability of a positive SARS-CoV-2 culture.
Probability computed by logistic regression of culture outcome as a function of viral RNA copy numbers detected by qRT-PCR in samples from seronegative volunteers. Shaded area is the approximate 95% confidence interval from the regression. **A.** Mid-turbinate swab. **B.** Saliva.

### References

1. Vogels CBF, Watkins AE, Harden CA, et al. SalivaDirect: A simplified and flexible platform to enhance SARS-CoV-2 testing capacity. Med (N Y) **2021**; 2:263-280.e6.

2. Matsuyama S, Nao N, Shirato K, et al. Enhanced isolation of SARS-CoV-2 by TMPRSS2-expressing cells. PNAS **2020**; 117:7001–7003.

3. Stadlbauer D, Amanat F, Chromikova V, et al. SARS-CoV-2 Seroconversion in Humans: A Detailed Protocol for a Serological Assay, Antigen Production, and Test Setup. Curr Protoc Microbiol **2020**; 57:e100.

4. Wrapp D, Wang N, Corbett KS, et al. Cryo-EM structure of the 2019-nCoV spike in the prefusion conformation. Science **2020**; 367:1260–1263.

5. Mullins KE, Merrill V, Ward M, et al. Validation of COVID-19 serologic tests and large scale screening of asymptomatic healthcare workers. Clin Biochem **2021**; 90:23–27.

6. Klein MN, Wang EW, Zimand P, et al. Kinetics of SARS-CoV-2 antibody responses pre-COVID-19 and post-COVID-19 convalescent plasma transfusion in patients with severe respiratory failure: an observational case-control study. J Clin Pathol **2021**; :jclinpath-2020-207356.

7. Li T, Chung HK, Pireku PK, et al. Rapid High-Throughput Whole-Genome Sequencing of SARS-CoV-2 by Using One-Step Reverse Transcription-PCR Amplification with an Integrated Microfluidic System and Next-Generation Sequencing. J Clin Microbiol **2021**; 59:e02784-20.

8. Genome Mapping Pipeline ngs_mapper. Viral Diseases Branch - WRAIR/NMRC, 2020. Available at: https://github.com/VDBWRAIR/ngs_mapper. Accessed 18 July 2021.

9. O'Toole Á, Hill V, McCrone JT, Scher E, Rambaut A. cov-lineages/pangolin. CoV-lineages, 2021. Available at: https://github.com/cov-lineages/pangolin. Accessed 15 July 2021.

10. Trevor Bedford, Richard Nehe, James Hadfield, et al. Nextstrain SARS-CoV-2 resources. 2020. Available at: https://nextstrain.org/sars-cov-2/. Accessed 29 July 2021.

11. Li H. Aligning sequence reads, clone sequences and assembly contigs with BWA-MEM. arXiv:13033997 [q-bio] **2013**; Available at: http://arxiv.org/abs/1303.3997. Accessed 29 July 2021.

12. Thorvaldsdóttir H, Robinson JT, Mesirov JP. Integrative Genomics Viewer (IGV): high-performance genomics data visualization and exploration. Brief Bioinform **2013**; 14:178–192.

13. Danecek P, Bonfield JK, Liddle J, et al. Twelve years of SAMtools and BCFtools. Gigascience **2021**; 10:giab008.

14. Wilm A, Aw PPK, Bertrand D, et al. LoFreq: a sequence-quality aware, ultra-sensitive variant caller for uncovering cell-population heterogeneity from high-throughput sequencing datasets. Nucleic Acids Res **2012**; 40:11189–11201.
